# Simulating the Impacts of Interregional Mobility Restriction on the Spatial Spread of COVID-19 in Japan

**DOI:** 10.1101/2020.12.28.20248926

**Authors:** Keisuke Kondo

## Abstract

This study develops a spatial Susceptible–Exposed–Infectious–Recovered (SEIR) model that analyzes the effect of interregional mobility on the spatial spread of the coronavirus disease 2019 (COVID-19) outbreak in Japan. National and local governments have requested that residents refrain from traveling between 47 prefectures during the state of emergency. However, the extent to which restricting the interregional mobility prevents infection expansion has not been elucidated. Our spatial SEIR model describes the spatial spread pattern of COVID-19 when people commute to a prefecture where they work or study during the daytime and return to their residential prefecture at night. We assume that people are exposed to infection risk during their daytime activities. According to our simulation results, interregional mobility restriction can prevent geographical expansion of the infection. However, in prefectures with many infectious individuals, residents are exposed to higher infection risk when their mobility is restricted. Our simulation results also show that interregional mobility restriction plays a limited role in reducing the national total number of infected individuals.

## 1. Introduction

Non-pharmaceutical interventions (NPIs), which are public health measures for the prevention and control of infection, have played a key role in combating the coronavirus disease 2019 (COVID-19) outbreak.^1–3^ To reduce the spatial spread of severe acute respiratory syndrome coronavirus 2 (SARS-CoV-2), national and local governments have enforced not only personal NPIs such as hand washing and mask-wearing, but also strict NPIs such as restricted movements, events, and travel, school closures, and quarantine.^4,5^ Among the strictest NPIs are lockdowns, which restrict our social and economic activities and trigger severe economic downturns.^2^

National and local governments in Japan have requested that residents refrain from traveling between 47 prefectures during the state of emergency, which was declared on April 7 of 2020.^5,6^ Although the interregional mobility restriction can be relaxed to include some outings after the state of emergency, the low level of lockdown restriction (i.e., voluntary compliance without penalties) was the first such experience in Japan.^7,8^ Therefore, how interregional mobility restriction limits the expansion of infection in Japan’s context is largely unknown, although many previous studies have emphasized the importance of understanding the spatial dynamics of spread.^9–16^ Effective control measures that prevent spatial spread of SARS-CoV-2 are urgently demanded, and how NPIs such as travel restrictions and social distancing mitigate the epidemic must be investigated.^17–27^ The present study aims to provide meaningful implications for combating the COVID-19 pandemic through interregional mobility restrictions.

To analyze how interregional mobility affects the spatial spread of SARS-CoV-2, we developed a spatial Susceptible–Exposed–Infectious–Recovered (SEIR) model. As the virus spreads through face-to-face contact, a spatial network of contagion was built by introducing interregional mobility into the standard SEIR model. The model assumes that people commute to a region of work or study in the daytime and return to their residential region at night. It further assumes that people are exposed to SARS-CoV-2 infection risk during their daytime activities, meaning that residents in one region are exposed to heterogeneous infection risks of SARS-CoV-2.

To simplify the spatial network analysis, the interregional mobility is mathematically treated as an origin–destination (OD) matrix. We demonstrate that our spatial SEIR model reduces to the standard SEIR model when the off-diagonal elements of the OD matrix are zero (i.e., when people remain in their residential regions). Therefore, our spatial SEIR model can be viewed as a generalized version of the standard SEIR model.

The developed approach can be also viewed as a spatial version of the social interaction approach. The social interaction approach constructs a contact matrix across various classes, such as age and gender.^19,25,27–29^ Similarly, face-to-face interactions across different regions are incorporated into the OD matrix.

The daily OD matrix is constructed from the interregional mobility data obtained by geospatial information technology, namely, from the locational information of mobile phone users. These data capture the specific situations of individuals, such as commuting to work on weekdays and remaining in the residential region or traveling to another region during the weekends. By tracking the interregional mobility on each month, day, and time of day throughout one year, we successfully captured the daily interregional mobility flows in the counterfactual situation.

This study aims to implicate effective control measures based on a simulation analysis. Because mitigating the COVID-19 pandemic is an urgent priority, an epidemic model that guides the planning of efficient control measures is essential when few ideal data are available.^30–32^ The spatial SEIR model assumes that the past interregional mobility trend will continue in future, regardless of how the COVID-19 pandemic evolves. Comparing those simulated from the SEIR model under the free mobility assumption with those simulated under the strict interregional mobility restriction, this study evaluates how restricting movement mitigates the spatial infection spread.

## 2. Methods

### 2.1. SEIR model without Interregional Mobility

In the absence of interregional mobility, the SEIR model assumes that infection in one region is independent of infection in all other regions. In other words, infection expansion in one region does not affect the infection dynamics in other regions. Later, this baseline model will incorporate interregional mobility as a source of the spatial infection expansion.

Suppose that there are *m* regions and that region *i* has population *N*_*i*_. The total national population is expressed as 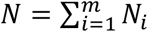. The birth and death rates are excluded from the model, meaning that the national total population is fixed over time. The regional population distribution is also fixed because residential change is forbidden in this model. The daily mobility (e.g., commuting and travel) is similarly forbidden in the baseline SEIR model, but this assumption will be relaxed later.

Let *S*_*i*_ (*t*), *I*_*i*_ (*t*), and *R*_*i*_ (*t*) denote the number of susceptible, infectious, and recovered residents, respectively, in region *i* at date *t*. Let *E*_*i*_ (*t*) denote the number of residents in region *i* at date *t* who have been exposed to the COVID-19 infection, but who are in the latent period and not yet infectious. In the SEIR model without a spatial network, the epidemic dynamics are expressed as follows:

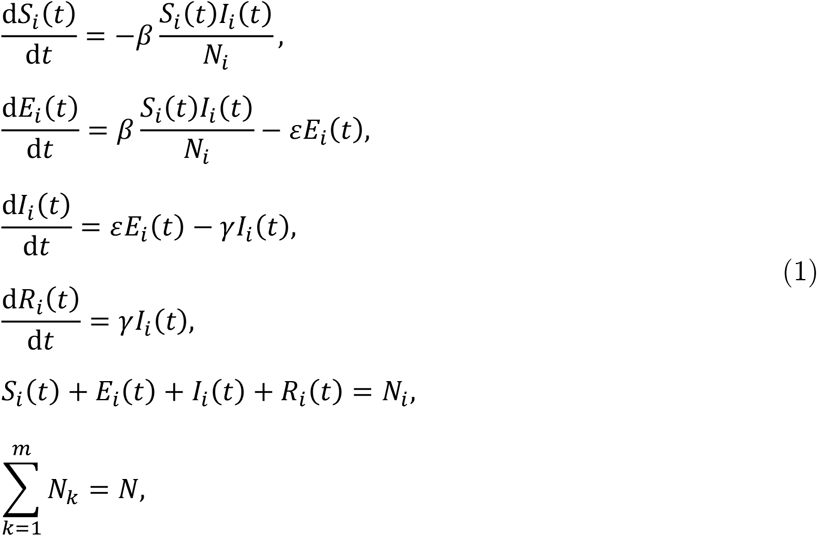

where *β* is the transmission rate parameter, *ε* is the incubation rate, and *γ* is the recovery rate.

The SEIR model without interregional mobility gives the baseline results for comparison with the extended model. From this comparison, we can observe how interregional mobility changes the results. A main distinguishing feature of the spatial SEIR model is the force of infection *λ*(*t*), which measures the rate at which susceptible individuals contract the infection. This term in the standard SEIR model *βI*_*i*_ (*t*)/*N*_*i*_ is independent of infection in all other regions. We highlight how relaxing the assumption of interregional mobility changes the specified force of infection.

### 2.2. SEIR model with Interregional Mobility

This study introduces interregional mobility as a spatial network of contagion into the standard SEIR model. The basic assumptions are those of the above-mentioned baseline setting. As an additional assumption, interregional mobility causes geographical infection expansion through person-to-person contact.

Individuals are assumed to be exposed to infection risk only in the region they occupy in the daytime. For example, suppose that an individual lives in one region and commutes to another region during the daytime. This interregional mobility provides two possible transmission channels: transfer of the disease from the region occupied by the commuting individual to the residential region and spread of the disease from the residential region to the region occupied by the commuting individual. Thus, interregional mobility spreads the infection disease across spatial domains.

The interregional mobility can be modeled by spatial network analysis. Let ***π***(*t*) denote the OD probability matrix across regions on date *t*:

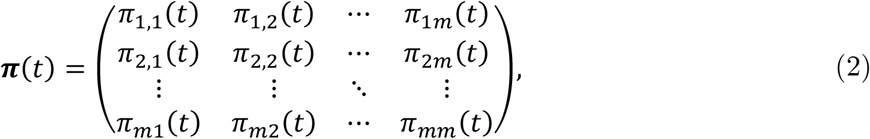

where *π*_*ij*_ (*t*) represents the individual’s probability of traveling from region *i* to region *j* on date *t*. The elements of ***π***(*t*) along a row must sum to one by the definition of probability. To simplify the calculation, we assume that this probability matrix is independent of the infection conditions over time (i.e., is not endogenous). However, the model admits exogenous seasonal variations or NPIs such as interregional mobility restriction.

In terms of the OD matrix, we can calculate the expected mobility flow from region *i* to region *j* and the expected daytime population in each region. The mobility flow from region *i* to region *j* is calculated as

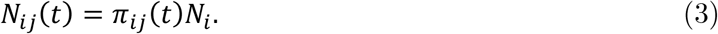

The expected total population in the daytime in region *i* is calculated as

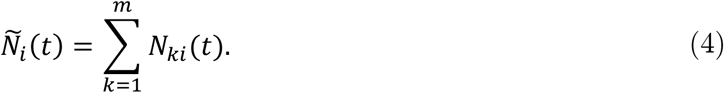

Similarly, the expected mobility flows of the susceptible, exposed, infectious, and recovered individuals are respectively expressed as *S*_*ij*_ (*t*) = *π*_*ij*_(*t*)*S*_*i*_ (*t*), *E*_*ij*_(*t*) = *π*_*ij*_ (*t*)*E*_*i*_(*t*), *I*_*ij*_(*t*) = *π*_*ij*_(*t*)*I*_*i*_ (*t*), and *R*_*ij*_ (*t*) = *π*_*ij*_(*t*)*R*_*i*_ (*t*). During the daytime, the expected numbers of susceptible, exposed, infectious, and recovered individuals are respectively expressed as 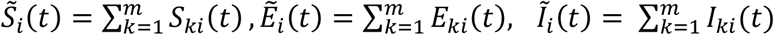 and 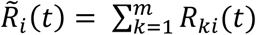. Note that the OD probability matrix is assumed to be common between the susceptible, exposed, infectious, and recovered individuals.

Importantly, the spatial distribution of infectious individuals in the daytime affects the infection risk in each region. During nighttime, the infectious individuals return to their residential regions. Therefore, we count cases of the infection within the residential regions. When residents in region *i* are exposed to heterogeneous infection risk in each region that they occupy during the daytime, the infection dynamics in region *i* is given by

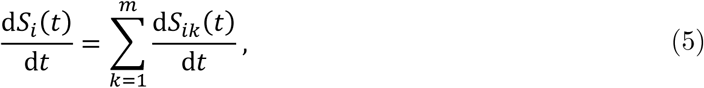

where d*S*_*ij*_ (*t*)/d*t* represents the transition that susceptible individuals residing in region *i* and staying in region *j* during the daytime become infected, and the sum of them in terms of region *j* represents the transition in the residential region *i*.

Consider residents in region *i* who remain in region *i*. The infection dynamics is expressed as follows:

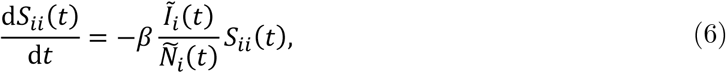

where the force of infection *βĨ*_*i*_ (*t*)/*Ñ*_*i*_ (*t*) depends on the number of infectious individuals and population in region *i* in the daytime. The infection dynamics for individuals who reside in region *i* and stay in region *j* in the daytime is expressed as

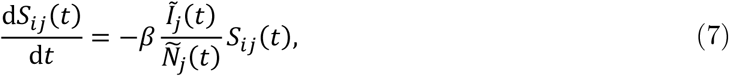

where the force of infection *βĨ*_*i*_ (*t*)/*Ñ*_*i*_ (*t*) depends on the number of infectious individuals and population in region *j* in the daytime. Thus, residents in region *i* are exposed to heterogeneous infection risks.

The overall transmission of infection in region *i* is expressed as the sum of the transmission of infection in terms of each outflow, which is given by

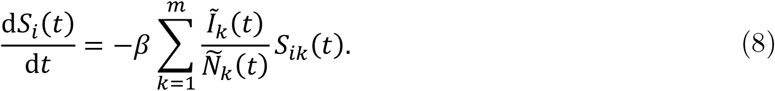

Finally, the dynamic system of equations of the spatial SEIR model with interregional mobility is given by

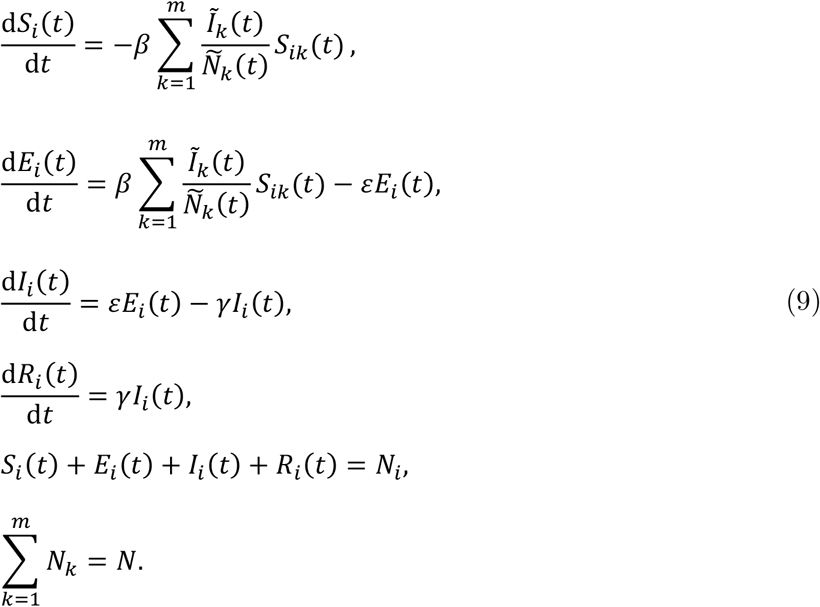

When the interregional mobility is restricted, the diagonal and off-diagonal elements of the OD matrix ***π***(*t*) take values 1 and 0, respectively. Under this assumption, we see that *N*_*ij*_ (*t*) = *S*_*ij*_ (*t*) = *E*_*ij*_ (*t*) = *I*_*ij*_ (*t*) = *R*_*ij*_ (*t*) = 0 for all *j*(≠ *i*) and that 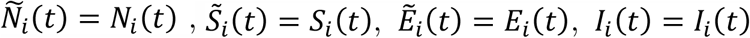, and 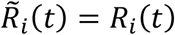. That is, the spatial SEIR model with interregional mobility reduces to the baseline SEIR model. The developed spatial SEIR model with interregional mobility can thus be viewed as a generalized version of the SEIR model.

### 2.3. Simulation Setting

The study objective was to evaluate how interregional mobility restrictions arrest the spatial spread of the COVID-19 infection. For this purpose, we compared the simulation results of the spatial SEIR model with and without interregional mobility. The difference between the two sets of results revealed the impact of interregional mobility on the spread of COVID-19 infection.

To uncover the condition under which restricting the interregional mobility effectively prevents spatial infection spread, we also considered another type of NPI implemented. The degree of the NPIs (scaling factor of transmission) is defined as *α*(*t*) ∈ (0, 1), and the time-varying transmission rate is given by *β*(*t*) = *α*(*t*)*β*. Note that we do not consider heterogeneous degree of the NPIs in each region, because we focus only on how the heterogeneous contact rates through the interregional mobility affect the spatial infection expansion.

Table 1 presents the parameters settings of the simulation analysis. We assume that average incubation and infectious periods *ℓ*_*ε*_ and *ℓ*_*γ*_ were assumed as 5 days and 10 days, respectively.^2,33–35,^ The infectiousness probability of an exposed individual was given as *ε* = 1/*ℓ*_*ε*_, and the recovery probability of an infected individual was given by *γ* = 1/*ℓ*_*γ*_. The transmission rate was determined as *β* = *R*_0_*γ* based on the standard SEIR model. The basic reproduction number *ℛ*_0_ was set to 2.6,^33^ close to that obtained by other studies in Japan.^36,37^ These parameter settings were common to all simulation scenarios except the intervention degree *α*(*t*).

**Table 1.** Parameter settings in the simulation. These parameter settings are common to each scenario.

| Parameters | Explanation | Value | References |
| --- | --- | --- | --- |
| $\mathcal{R}_0$ | Basic reproduction number | 2.6 | 36,38 |
| $\ell_\varepsilon$ | Average incubation period | 5 days | 2,31,33-35 |
| $\ell_\gamma$ | Average infection period | 10 days | 2,33 |

In this study, the degree of NPIs was exogenously given by the parameter *α*(*t*) as a future policy target parameter, and two case scenarios were simulated: modest convergence, and worsening cases. Figure 1 shows the intervention degree in both scenarios. Although the values were arbitrarily set for future scenarios, we attempted to predict their potential ranges from the estimated effective reproduction numbers.^38,31^ For example, when declaring a state of emergency on April 7 of 2020, the Government requested that person-to-person contact be reduced by 70% at least, meaning that the intervention degree in Figure 1 was 0.3. For the basic reproduction number *ℛ*_0_ in the baseline SEIR model, the intervention degree must be lower than 1/*ℛ*_0_ in the convergence scenario (i.e., the effective reproduction number must be lower than 1), and 1/*ℛ*_0_ or higher in most months of the worsening case scenario.

**Figure 1.**
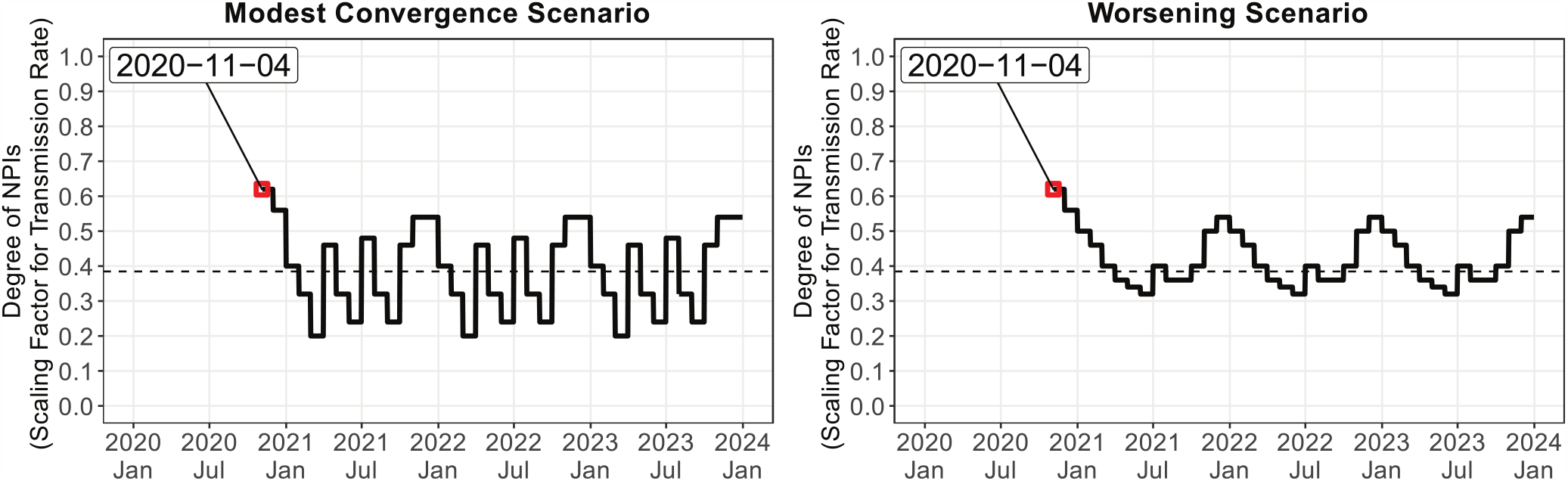
Simulation settings of the degree of the non-pharmeceutical interventions as a policy target parameter. Simulations were started on November 4, 2020. The dashed line indicates *α(t)* = l/ℛ_0_ in the baseline SEIR model.

The simulation was started on November 4 of 2020, the approximate date before the onset of the third wave of the pandemic. In the Supplemental Material, we provide additional simulation results. As a robustness check, we also evaluated how the starting date influenced the simulation results (e.g., April 7 of 2020 and August 17 of 2020). In addition, we evaluated the effect of the interregional mobility restriction only for Tokyo.

### 2.4. Data

Positive COVID-19 cases in each prefecture of Japan are reported by the prefectural government and the Ministry of Health, Labour and Welfare (MHLW). This study collected the daily and cumulative numbers of positive tests reported by each prefectural government until November 25 of 2020. The slight differences between the numbers reported by the prefectural governments and the MHLW did not affect the qualitative results.

Figure 2 is a snapshot of the geographical distribution of the numbers of positive cases on August 17 of 2020 and November 10 of 2020. The COVID-19 infection numbers were highest in Tokyo (161 and 293 patients testing positive, respectively), followed by Osaka. Although the infection tended to expand in prefectures with large cities, such as Tokyo, Osaka, Nagoya, Sapporo, Yokohama, and Saitama, other prefectures occasionally experienced a sudden rise in infection numbers.

**Figure 2.**
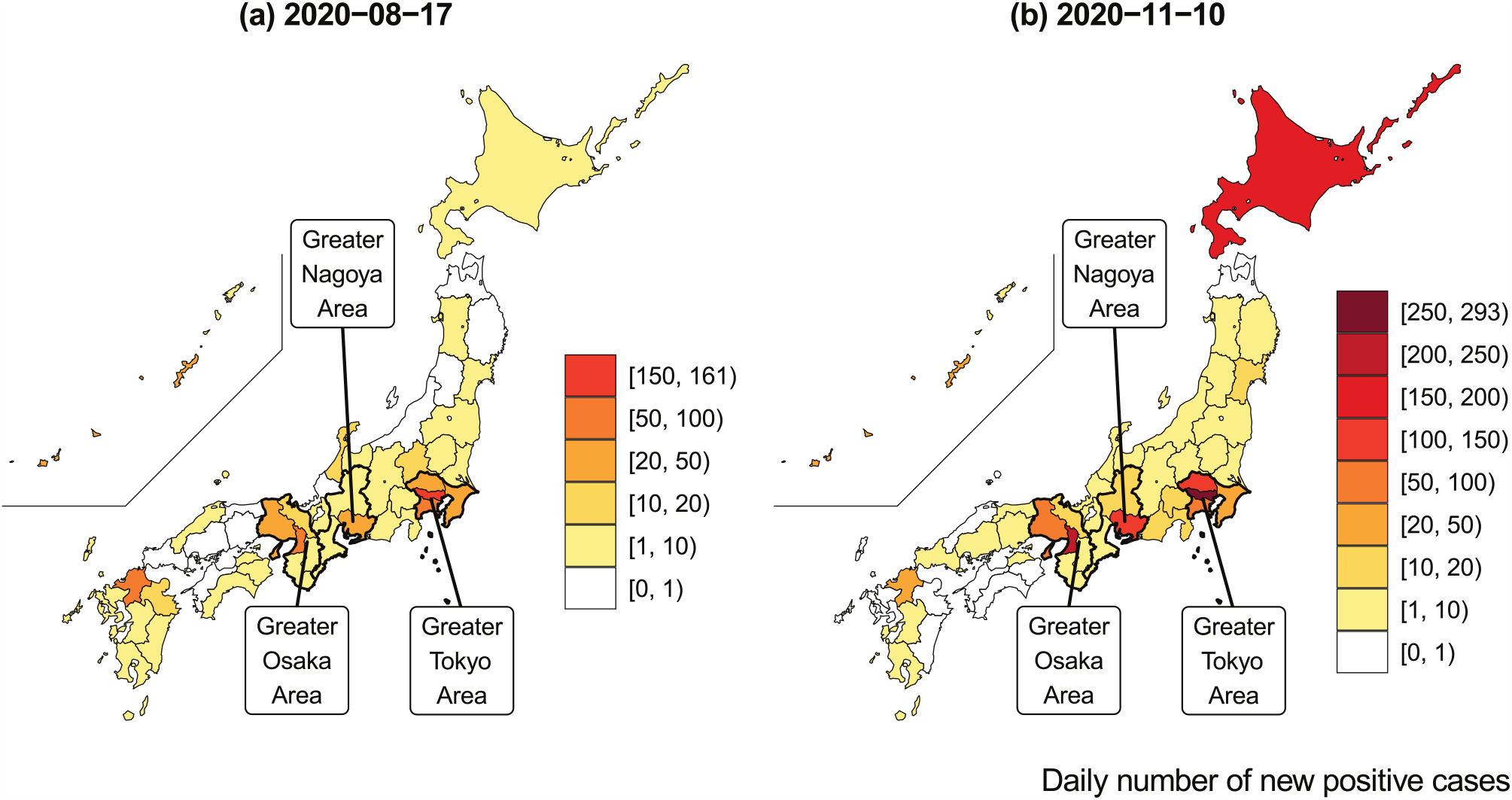
COVID-19 infection across prefectures. This map was constructed from of the daily number of positive tests reported by each prefectural government.

Considering the observed cumulative number of positive patients and the average number of days in the incubation and infectious periods (*ℓ*_*ε*_ and *ℓ*_*γ*_, respectively), the variables in the SEIR model were constructed as follows:

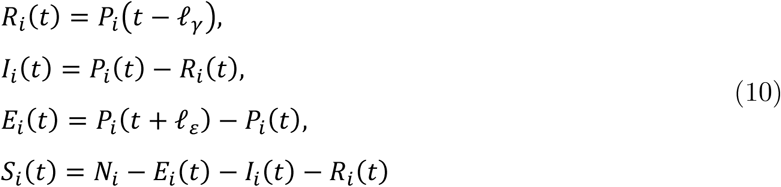

where *P*_*i*_ (*t*) represents the cumulative number of positive tests reported in prefecture *i* at date *t*. The total population in each prefecture *i* was based on October 2019 data and was fixed over time (i.e., no migration was assumed across prefectures).

These variables were calculated from the observed data before the simulation start date, and were predicted by the spatial SEIR model after the simulation start date.

The theoretical part of the model was based on the probabilistic mobility of individuals, but the OD probability matrix in the empirical part was estimated from the observed data on interregional mobility, which were derived from the locational information of mobile phone users. The Regional Economy and Society Analyzing System (RESAS), a web application developed by the Headquarters for Overcoming Population Decline and Vitalizing Local Economy in Japan at the Prime Minister’s Office, was released on April 21 of 2015.^39^ The RESAS app visualizes many types of data in Japan, including a dynamic map of the inter-municipal human flows (the From–To Analysis) based on the Mobile Spatial Statistics of NTT DOCOMO.^40^ The detailed information of inter-municipal flows are available by gender, age, year, month, day of the week (weekdays and weekends), and time of day (4 am, 10 am, 2 pm, 8 pm).

This study applied the monthly data of interregional flows from September of 2015 to August of 2016 by day of week (weekday or weekend). Although the interregional flow data is visualized on the RESAS app until 2020, the application programming interface can download the original data only within a restricted period (from September of 2015 to August of 2016). The daytime population in each prefecture was estimated from the inter-municipal flows observed at 2 pm. The inter-municipal flows were aggregated into inter-prefectural flows to match the observational unit of the COVID-19 infection data. In each scenario, the daily pattern of interregional mobility from September 2015 to August 2016 was assumed from the start date of the simulation to December 31 of 2023; that is, the same mobility pattern was repeated on the same day of each year.

From the data, we calculated the share of inter-prefectural mobility that matched the probability of interregional mobility. Let *C*(*t*) denote the OD matrix across the 47 prefectures on date *t*:

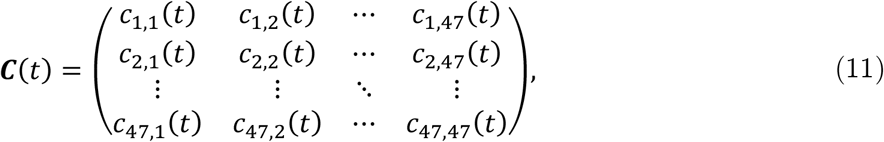

where *c*_*ij*_ (*t*) represents the mobility flow (i.e., number of people) from prefecture *i* to prefecture *j* on date *t*. This OD matrix was row-standardized to express the share as follows:

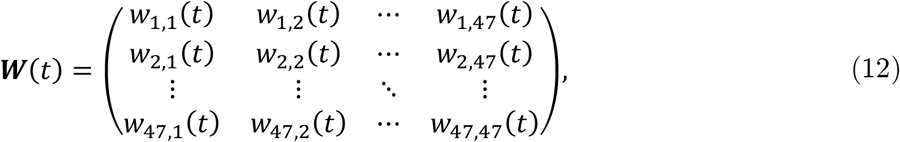

where *w*_*ij*_ (*t*) represents the share of residents in prefecture *i* who were staying in prefecture *j* on date *t*. The row-standardization of *C*(*t*) gives the share of inter-prefectural flows as follows:

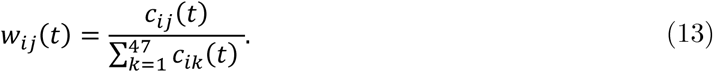

This weight matrix corresponds to the probabilistic OD matrix in the theoretical model.

Figure 3 shows the inter-prefectural OD matrices at 2 pm on weekdays and weekends in April of 2016. The color strengths in the tile plots represent the flow share. Panel (a) shows that most of the residents remained in their home prefectures. Panel (b) focuses on the OD matrix in the Greater Tokyo area (Saitama, Chiba, Tokyo, and Kanagawa). More than 10 percent of the residents in the neighboring prefectures of Tokyo were in Tokyo at 2 pm on the weekdays of April 2016, but residents tended to remain in their home prefectures on weekends. This mobility trend affected the inter- and intra-prefectural spread of the infection in the Greater Tokyo area. Panels (b) of Figure 3 shows the OD matrices in the Greater Osaka areas. Although the inflows into Osaka from neighboring prefectures were smaller than in the Greater Tokyo area, Osaka also attracted people from neighboring prefectures.

**Figure 3.**
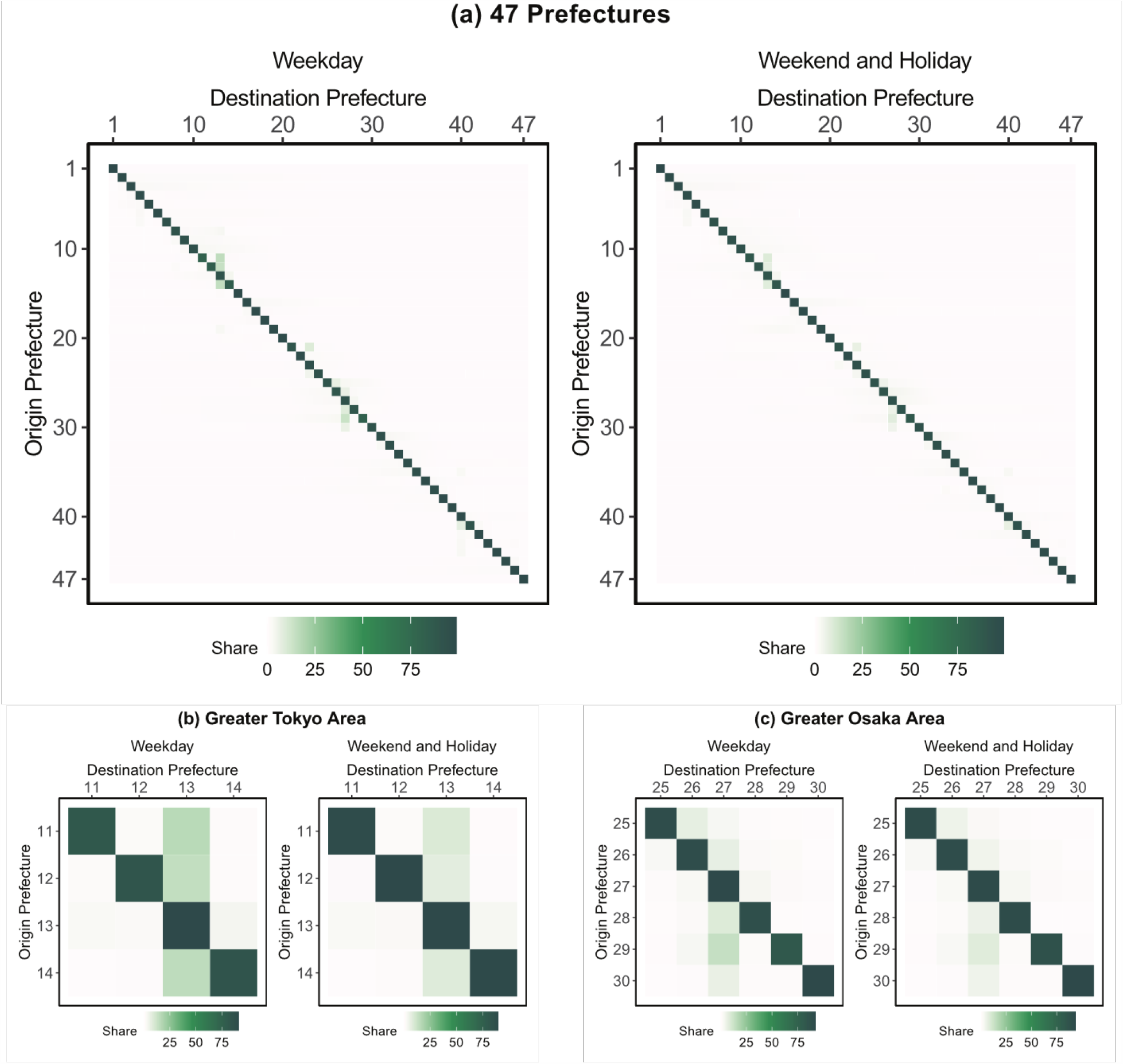
Origin-Destination matrix between prefectures, constructed from the interregional mobility data obtained by the “From-To Analysis” function of the RESAS app (supported by Mobile Spatial Statistics of NTT DOCOMO). The tile plot shows the OD flows at 2 pm on the weekdays of April, 2016.

Figure 4 shows the ratio of daytime and nighttime populations by day of the week (weekdays versus weekends). As shown in Panel (a), people residing in the neighboring prefectures of Tokyo and Osaka tended to concentrate in Tokyo and Osaka, respectively, during the daytime. However, Panel (b) of Figure 4 shows that the spatial distribution of the population diverged across the country on weekends. From the interregional mobility data based on the locational information of mobile phone users, we could precisely predict the spatial spread of infection through interregional commuting and travel on weekends, and the seasonal trend of the interregional mobility.

**Figure 4.**
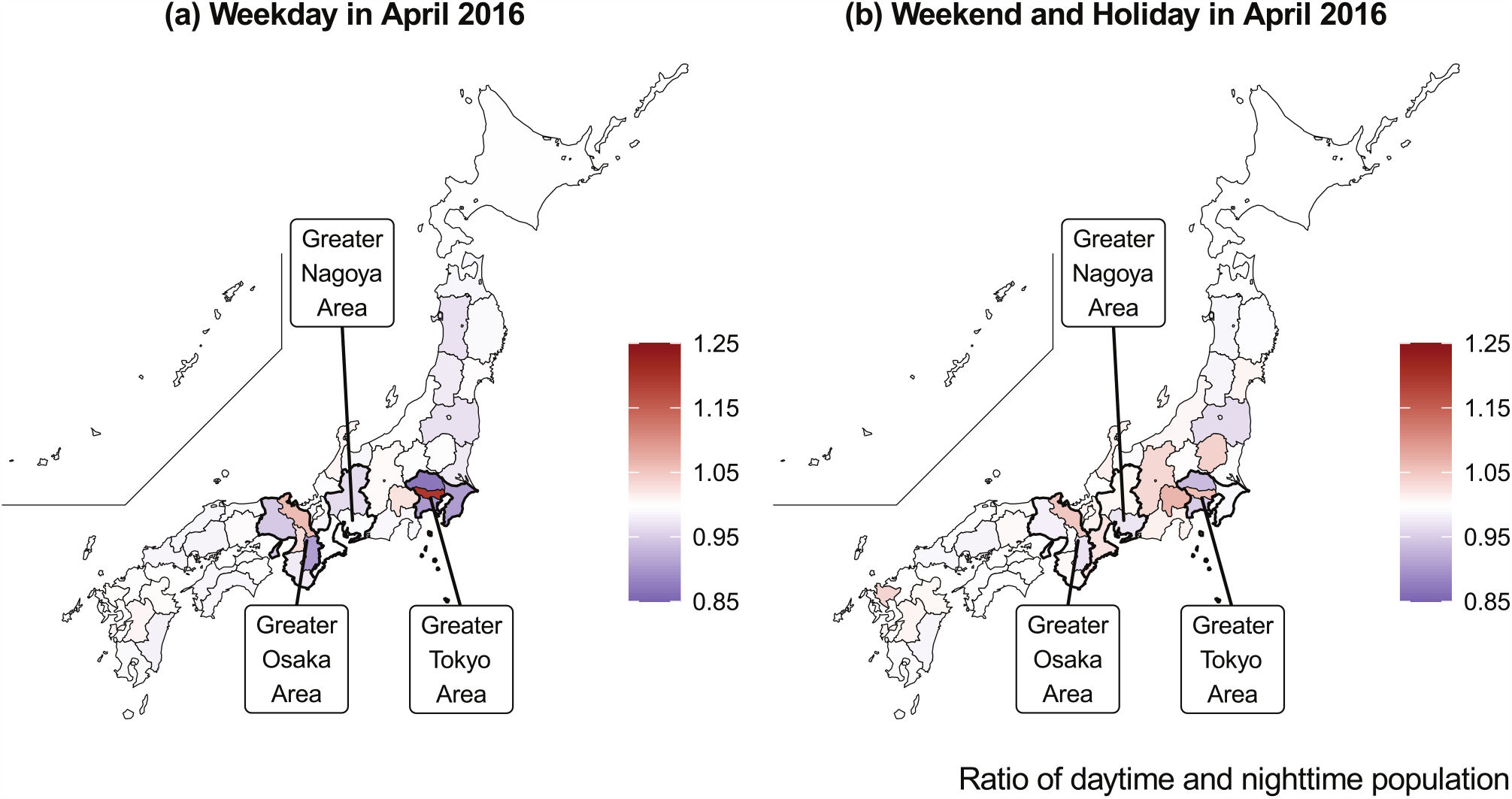
Ratio of daytime and nighttime population on weekday and weekend in April 2016, created from the interregional mobility data obtained by the “FromTo Analysis” function of the RESAS app (supported by Mobile Spatial Statistics of NTT DOCOMO)

## 3. Results

The simulations in this paper predicted the numbers of infectious individuals. All estimation results are available on the web application.

Figure 5 shows the simulation results in the modest convergence scenario. The simulated numbers of infectious individuals from the spatial SEIR model with and without interregional mobility (red and green lines, respectively), are compared with the observed number of infectious individuals (blue line).

**Figure 5.**
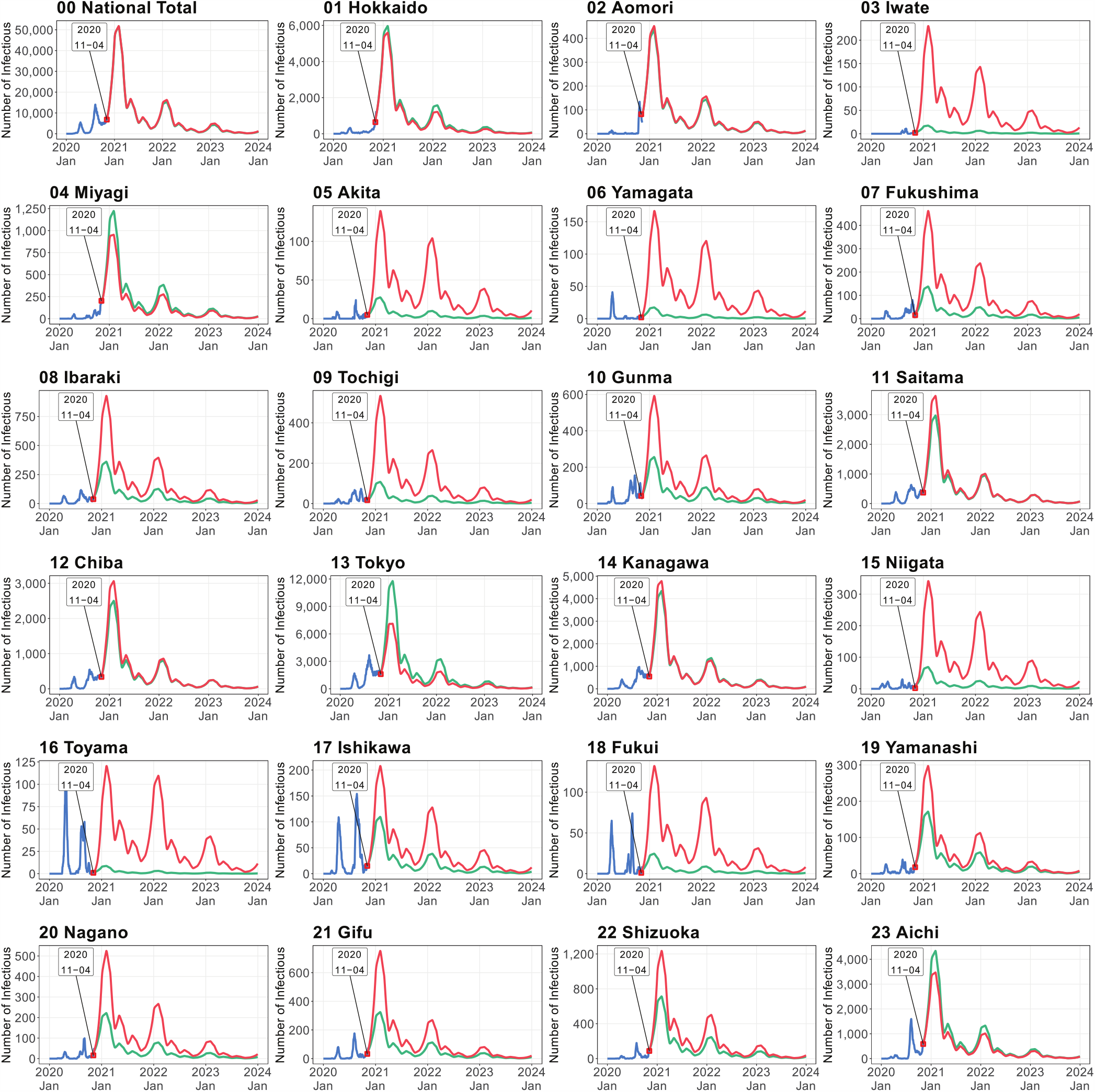
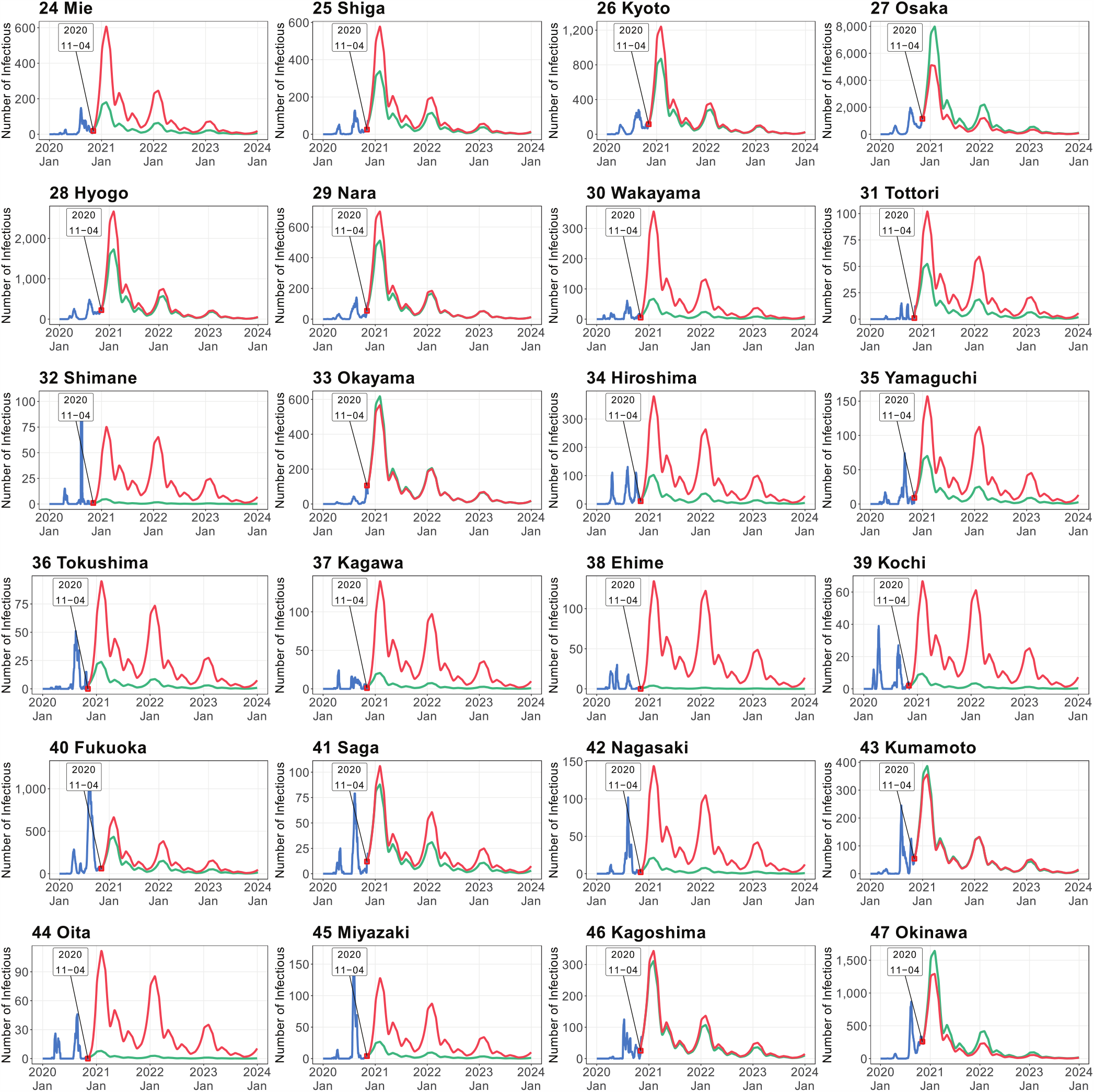
Simulated number of infectious persons by prefecture in modest convergence case scenario. Shown are the observed numbers of infectious individuals (blue lines), and the numbers of infectious individuals simulated by the spatial SEIR model with and without interregional mobility (red and green lines, respectively). Simulations were started on November 4 of 2020. Simulated number of infectious persons by prefecture in modest convergence case scenario

The gap between the red and green lines captures the influence of interregional mobility on the spatial infection spread of COVID-19. This gap tended to be large in rural areas with lower numbers of infectious individuals at the starting date of the simulation, such as Iwate, Akita, Yamagata, Fukushima, Ibaraki, Tochigi, Gunma, Niigata, Toyama, Fukui, Nagano, Gifu, Mie,

Wakayama, Shimane, Hiroshima, Yamaguchi, Tokushima, Kagawa, Ehime, Kochi, Nagasaki, Oita, and Miyazaki. When interregional mobility was included, the numbers of infectious individuals predicted by the spatial SEIR model were more than twice those predicted without interregional mobility, implying that interregional mobility caused the infection expiation in those prefectures.

In prefectures with large cities, such as Tokyo, Aichi, and Osaka, the spatial SEIR model with interregional mobility predicted lower numbers of infectious individuals than the model without interregional mobility. To understand why the interregional mobility generated these results, we calculated the ratios of the daytime and nighttime forces of infection in the Greater Tokyo, Nagoya, and Osaka areas.

Figure 6 plots the obtained ratios in the modest convergence case scenarios. Note that the ratio becomes one in the spatial SEIR model without interregional mobility, because the spatial population distributions during the daytime and nighttime are identical. As shown in Figure 6, the ratios in Tokyo, Aichi, and Osaka were lower than one and gradually converged to one.

**Figure 6.**
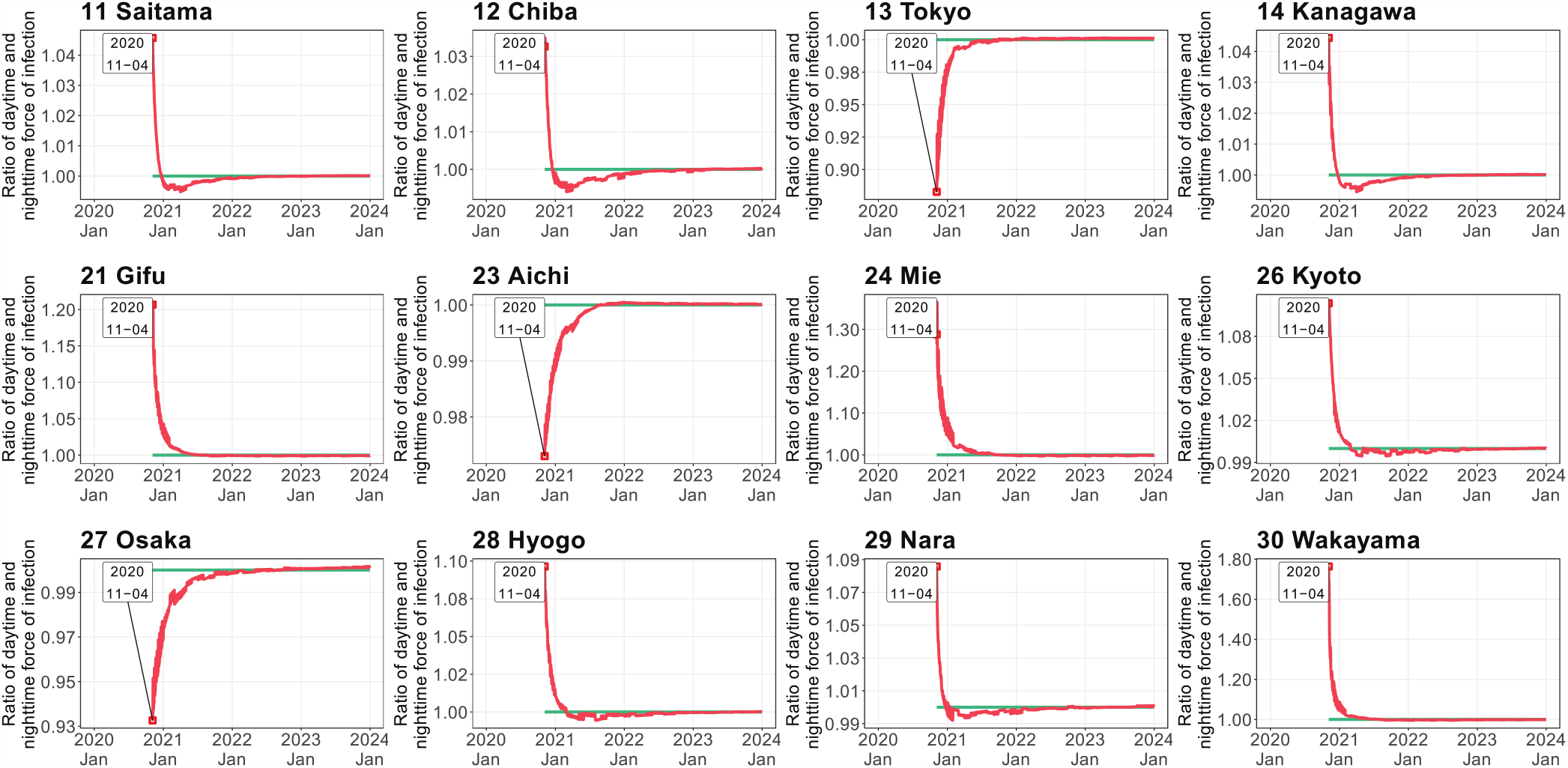
Ratio of daytime and nighttime force of infection in modest convergence case scenario (see caption of Figure 5 for details).

Although large cities generally attract more external people than smaller cities and towns, the force of infection during the daytime decreased because the influx from neighboring prefectures contained susceptible individuals. The outflux of infectious individuals from Tokyo, Aichi, and Osaka also decreased the daytime force of infection in those cities. That is, the ratios in the neighboring prefectures of Tokyo, Aichi, and Osaka were higher than one and gradually converged to one. Therefore, susceptible residents in Tokyo, Aichi, and Osaka were exposed to lower infection risk in the daytime than in the nighttime. Continuous interregional mobility gradually equalized the daytime and nighttime forces of infections within the Greater Tokyo, Nagoya, and Osaka areas.

Importantly, the national total numbers of infectious individuals were almost identical in the SEIR models with and without interregional mobility although those simulated from the SEIR without interregional mobility were always lower than those simulated from the SEIR with interregional mobility. In other words, restricting the interregional mobility slowed the speed of the infection spread but did not effectively reduce the infection expansion in the modest convergence scenario.

Figure 7 shows the simulation results in the worsening case scenario. Relative to the modest convergence scenario, restricting the interregional mobility widened the gap between the national total numbers of infectious individuals predicted by the SEIR models in the long term. This finding implies that restricting the interregional mobility lowered the growth rate of numbers of infectious individuals when the effective reproduction number exceeded one.

**Figure 7.**
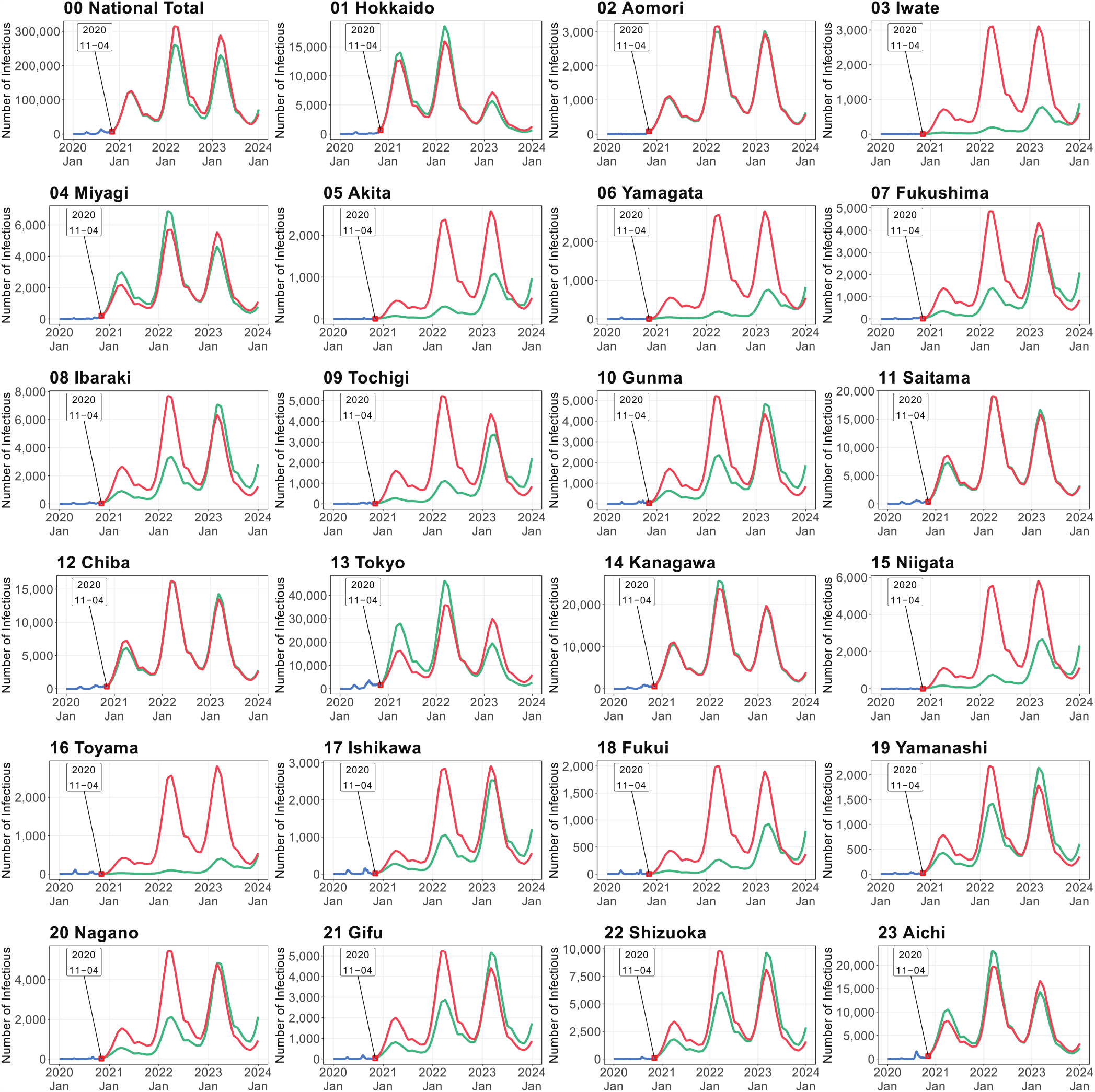
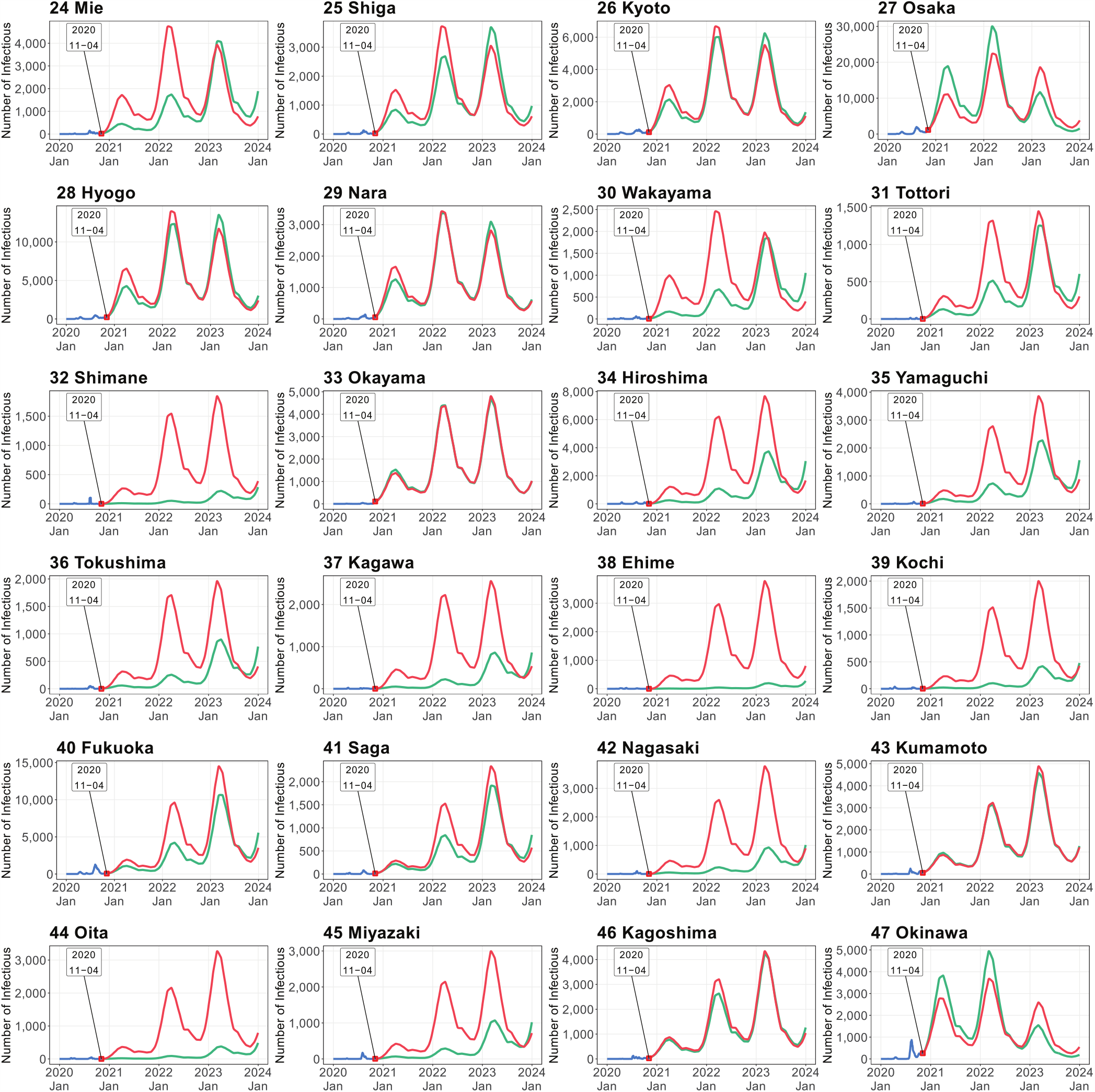
Simulated number of infectious persons by prefecture in worsening case scenario (see caption of Figure 5 for details).

Another important result was that interregional mobility accelerated the expansion of the COVID-19 infection in the short term in the rural areas, such as Iwate, Akita, Yamagata, Fukushima, Ibaraki, Tochigi, Gunma, Niigata, Toyama, Fukui, Nagano, Gifu, Mie, Wakayama, Shimane, Hiroshima, Yamaguchi, Tokushima, Kagawa, Ehime, Kochi, Nagasaki, Oita, and Miyazaki. On the other hand, there were prefectures that had relatively small gaps in number of infectious individuals between the SEIR models with and without interregional mobility, such as, Aomori, Okayama, Kumamoto, and Kagoshima.

## 4. Discussion

Our spatial SEIR model with interregional mobility revealed the effect of interregional mobility on the geographical infection spread of COVID-19. The simulation relies on the past interregional mobility data obtained from the locational information of mobile phone users. By analyzing these data, we could evaluate the differences in interregional mobility by season, day of the week, and period of the day. The government of Japan has recognized the effectiveness of high-frequency and real-time mobility data in mitigating the COVID-19 pandemic.^41^ In this study, the interregional mobility data were constructed into a daily OD matrix, showing the applicability of recent geospatial information technology in epidemic models.

From the simulation results of both scenarios, we can understand two important aspects of control measures. The first aspect involves the spatial spread of COVID-19. As expected, restricting the interregional mobility prevented the wide geographical spreading of the infection. This control measure is especially important for rural regions with scarce healthcare resources. Consistent with our simulation results, college student travel during the spring break has contributed to local infection transmission in the U.S.^21^

Surprisingly, in prefectures with large cities that attract outside workers (such as Tokyo and Osaka), the number of infections increased after restricting the interregional mobility. Although restricting mobility has reduced the total number of COVID-19 cases per capita in some U.S. cities,^20^ our simulation results from the spatial SEIR model suggest that the interregional mobility restriction has heterogeneous impacts on the infection expansion across regions. For example, an influx of uninfected persons from outside, and an outflux of infectious persons from regions with many infections, such as Tokyo and Osaka, will reduce the infection risk in the daytime in those regions. Therefore, restricting the interregional mobility without restricting intra-regional mobility will result in an increase in the infection risk to residents in large cities. To prevent further expansion of the infection in regions with many infections, additional strong NPIs should accompany the restrictions on interregional mobility. For example, contact restriction may be an effective control measure.^17^

The second aspect of control measures is whether the interregional mobility restriction can reduce the national total number of infections. Distinguishing between the short- and long term control measures is important, and the fundamental goal should be toward reducing the overall epidemic size.^18^

According to our simulation results, restricting the interregional mobility had a limited effect on reducing the national total number of infectious individuals. Imposing the mobility restriction cut the growth rate in national total number of infectious individuals only when the effective reproduction number was one or higher. Moreover, it was pointed out that regional lockdowns effectively reduced the overall epidemic size only when the transmission rate remained persistently low.^18^ These results imply that the efficacy of imposing the mobility restriction (in terms of reducing the overall epidemic size) is very sensitive to the timing of the restriction. In the Supplemental Material, we also evaluated other case scenarios (e.g., the effects of the timing of the mobility restriction and regional lockdown in Tokyo).

In conclusion, the interregional mobility restriction dominantly affected the spatial pattern and the speed of the infection spread and played a limited role in reducing the national total number of infections. The most important implication of restricting the interregional mobility is the avoidance of an epidemic peak that overwhelms the existing healthcare services, especially in rural regions where healthcare resources are typically scarce in Japan.^42^ In that sense, the mobility restriction is an effective control measure.

## Data Availability

The R code and data for this study are available on GitHub.
(https://github.com/keisukekondokk/spatial-seir)
The numerical simulation results in this study are provided on the web app "COVID-19 Simulator in Japan.”
(https://keisuke-kondo.shinyapps.io/covid19-simulator-japan/)

https://github.com/keisukekondokk/spatial-seir

https://keisuke-kondo.shinyapps.io/covid19-simulator-japan/

## Acknowledgements

I thank Masayuki Morikawa and Yoichi Sekizawa for their helpful comments and suggestions. Naturally, any remaining errors are my own. This research was conducted under the project at RIETI.

## Supplementary information

The R code and data for this study are available on GitHub. (https://github.com/keisukekondokk/spatial-seir)

The numerical simulation results in this study are provided on the web app “COVID-19 Simulator in Japan.” (https://keisuke-kondo.shinyapps.io/covid19-simulator-japan/)

