## Supplementary Material for "Simulating the Impacts of Interregional Mobility Restriction on the Spatial Spread of COVID-19 in Japan"

Keisuke Kondo*

RIETI

This online appendix provides simulation results for the number of infectious persons in the six case scenarios. The fourth and fifth cases were discussed in the main text.

*Research Institute of Economy, Trade and Industry (RIETI). 1-3-1 Kasumigaseki, Chiyoda-ku, Tokyo, 100–8901, Japan..

#### Contents

**- Online Appendix A.**

COVID-19 Simulator 3

**- Online Appendix B.**

Case 1: Rapid covergence case scenario at the early stage with time-constant transmission rate from April 7 of 2020 6

**- Online Appendix C.**

Case 2: Modest convergence case scenario with time-varying transmission rate from April 7 of 2020 10

**- Online Appendix D.**

Case 3: Modest convergence case scenario with time-varying transmission rate from August 17 of 2020 14

**- Online Appendix E.**

Case 4: Modest convergence case scenario with time-varying transmission rate from November 4 of 2020 18

**- Online Appendix F.**

Case 5: Worsening case scenario with time-varying transmission rate from November 4 of 2020 22

**- Online Appendix G.**

Case 6: Mobility restriction only for Tokyo and modest convergence case scenario with time-varying transmission rate from November 4 of 2020 26

### Online Appendix A.
#### COVID-19 Simulator

This study developed a Shiny application to visualize simulated numbers of the susceptible, exposed, infectious, and recovered individuals in each case scenario. All simulation results are provided on the web application.
(URL: `https://keisuke-kondo.shinyapps.io/covid19-simulator-japan/`).

Figure A.1 shows the top page of the web app. Users can select one of the six scenarios on the side bar. In the main panel, users can access original data and simulation results. For example, Figure A.2 shows the inter-prefectural flows on the web application (Click Spatial Network Data on the Visualization menu). The width of the line and the strength of the line color represent the size of flow. The map shows the share of people residing in each prefecture who stayed in Tokyo at 2 pm on weekday in April 2016. For example, 15–20 % of people residing in Saitama stayed in Tokyo at 2 pm on weekday in April 2016. All bilateral flows across the 47 prefectures are visualized on the web application.

[Figures A.1 and A.2]

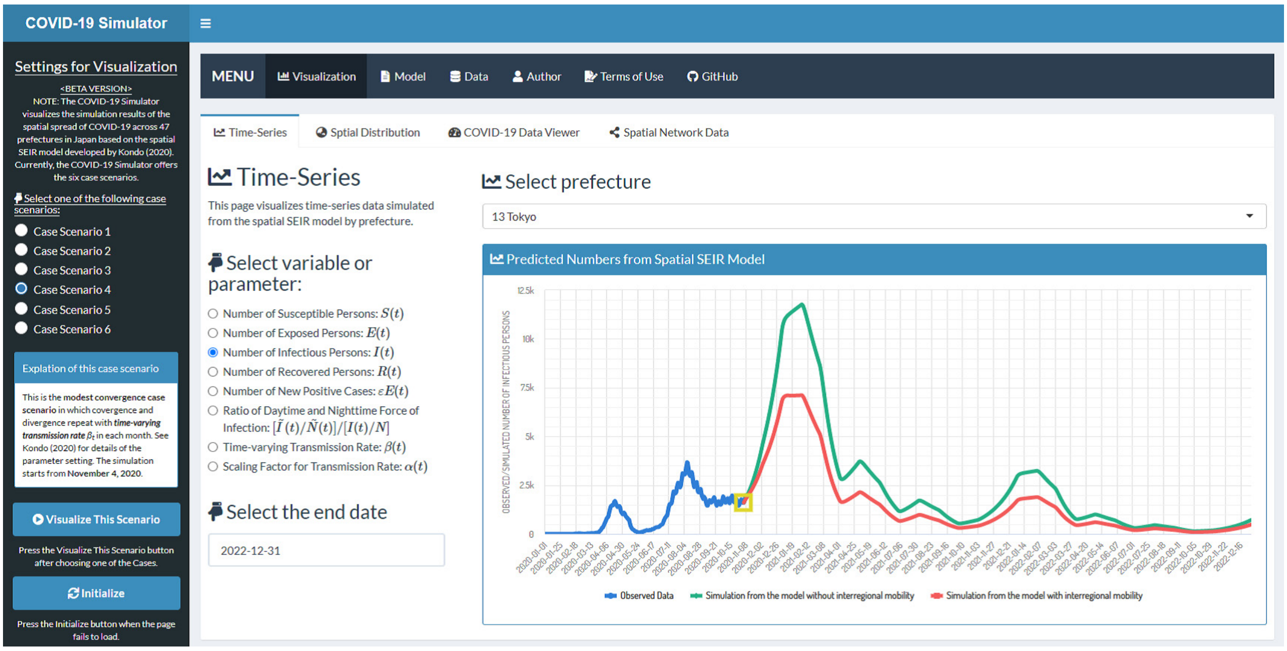


**Figure A.1.** COVID-19 Simulator

URL: `https://keisuke-kondo.shinyapps.io/covid19-simulator-japan/`

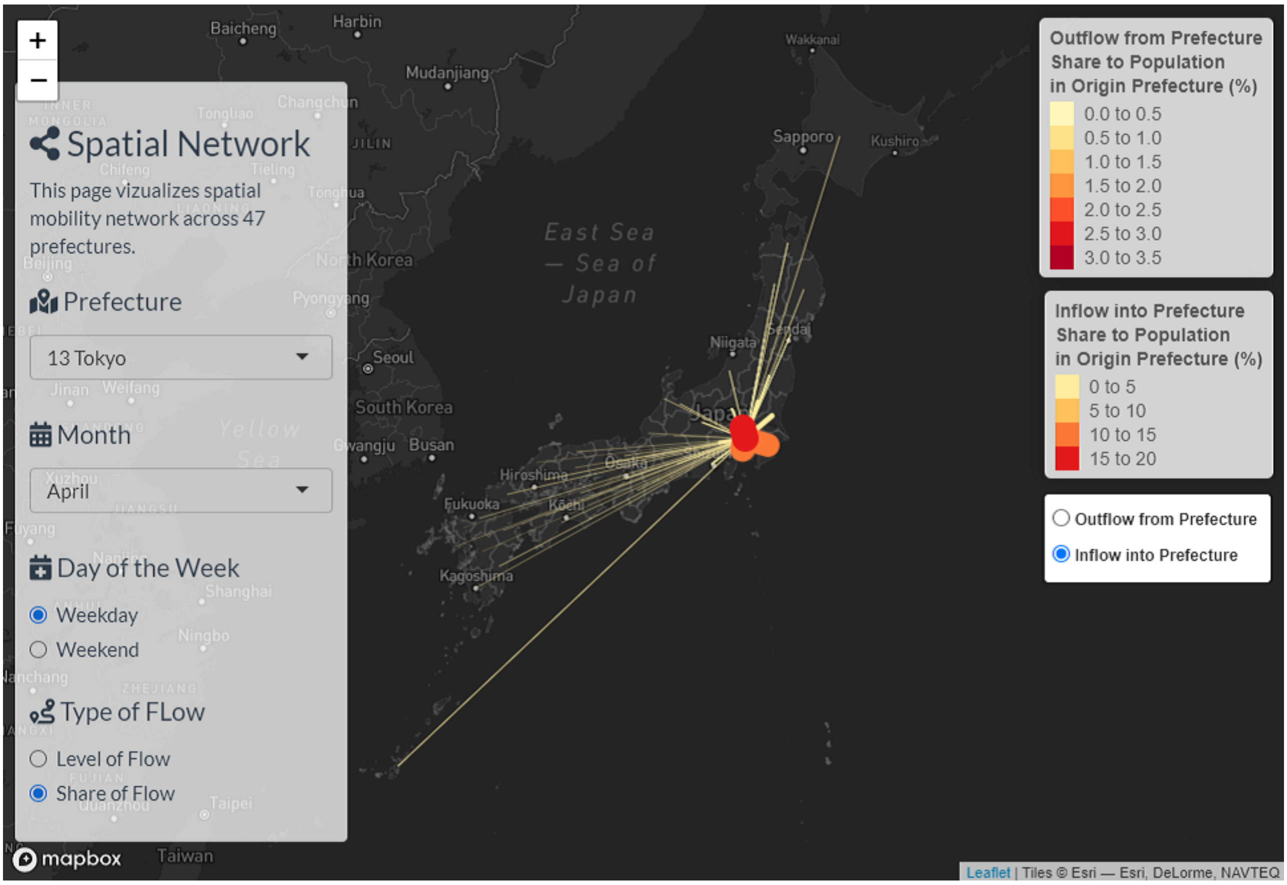


**Figure A.2.** Interregional Mobility across the 47 Prefectures

Note: See caption of Figure 4 for details. The map shows the share of people residing in each prefecture who stayed in Tokyo at 2 pm on weekday in April 2016. For example, 15–20 % of people residing in Saitama stayed in Tokyo at 2 pm on weekday in April 2016.

### Online Appendix B.

#### Case 1: Rapid covergence case scenario at the early stage with time-constant transmission rate from April 7 of 2020

Figure B.1 shows the simulation results in the rapid convergence case scenario, which was originally considered at the early stage before the declaration of the state of emergency on April 7 of 2020.

Table B.1 presents the specific values of intervention degree $\alpha(ts)$ used in the simulation. The transmission rate is time-constant so that the basic reproduction number is 0.75. The government of Japan requested at least to prevent the person-to-person contact by 70 % when declaring the state of emergency. In this situation, the basic reproduction number was assumed to be 2.5, and the government's request meant the effective reproduction number was 0.75.

[Table B.1 and Figure B.1]

**Table B.1.** Parameter Setting of Intervention Degree in Case Scenario 1

| Year | Month 1 | 2 | 3 | 4 | 5 | 6 | 7 | 8 | 9 | 10 | 11 | 12 |
|---|---|---|---|---|---|---|---|---|---|---|---|---|
| | Case 1 (Starting date of simulation: April 7 of 2020) | | | | | | | | | | | |
| 2020 | - | - | - | 0.2885 | 0.2885 | 0.2885 | 0.2885 | 0.2885 | 0.2885 | 0.2885 | 0.2885 | 0.2885 |
| 2021 | 0.2885 | 0.2885 | 0.2885 | 0.2885 | 0.2885 | 0.2885 | 0.2885 | 0.2885 | 0.2885 | 0.2885 | 0.2885 | 0.2885 |
| 2022 | 0.2885 | 0.2885 | 0.2885 | 0.2885 | 0.2885 | 0.2885 | 0.2885 | 0.2885 | 0.2885 | 0.2885 | 0.2885 | 0.2885 |
| 2023 | 0.2885 | 0.2885 | 0.2885 | 0.2885 | 0.2885 | 0.2885 | 0.2885 | 0.2885 | 0.2885 | 0.2885 | 0.2885 | 0.2885 |

Note: See caption of Table 2 for details.

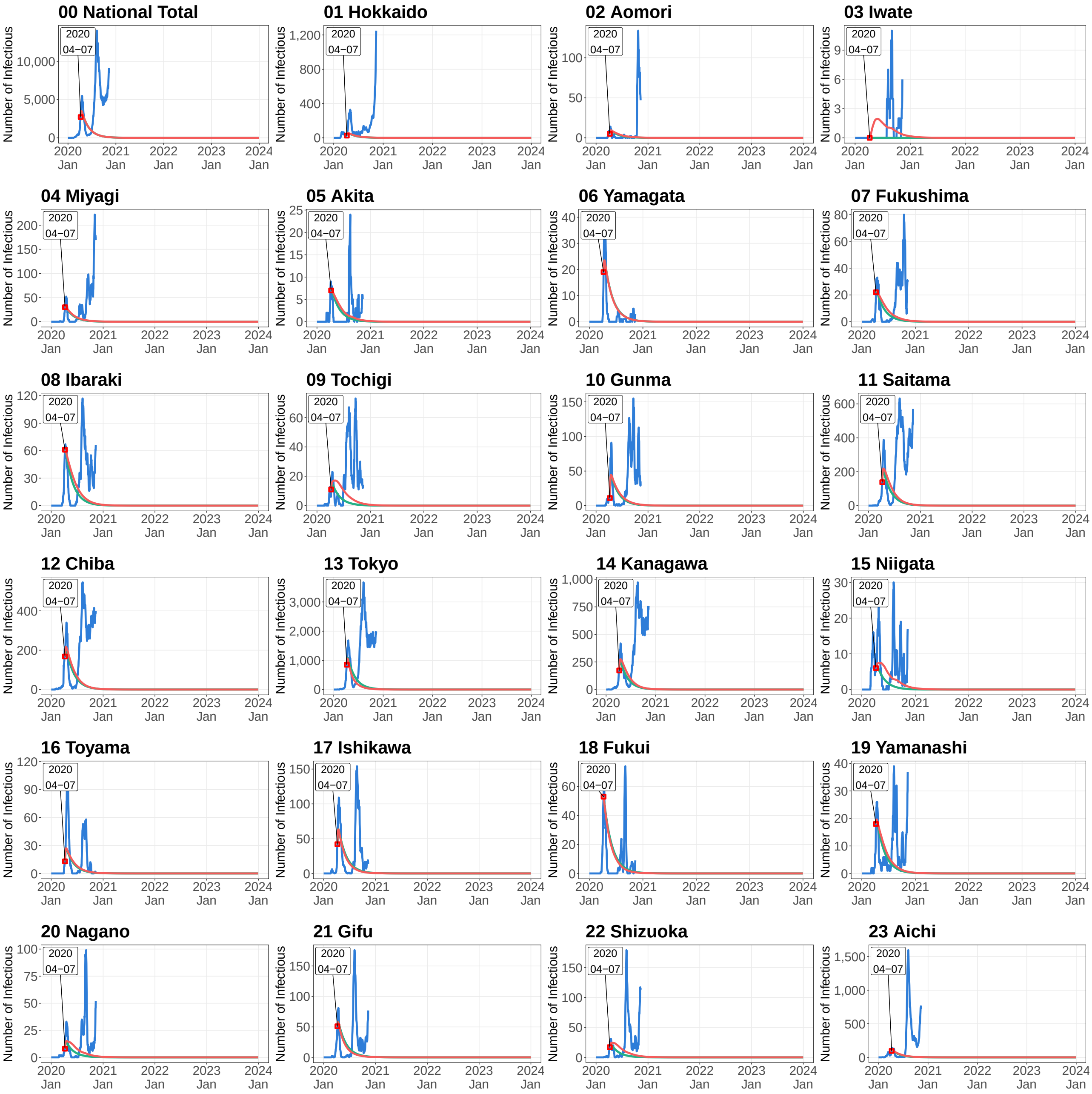


**Figure B.1.** Simulated Number of Infectious Persons by Prefecture in Case Scenario 1

Note: See caption of Figure 4 for details.

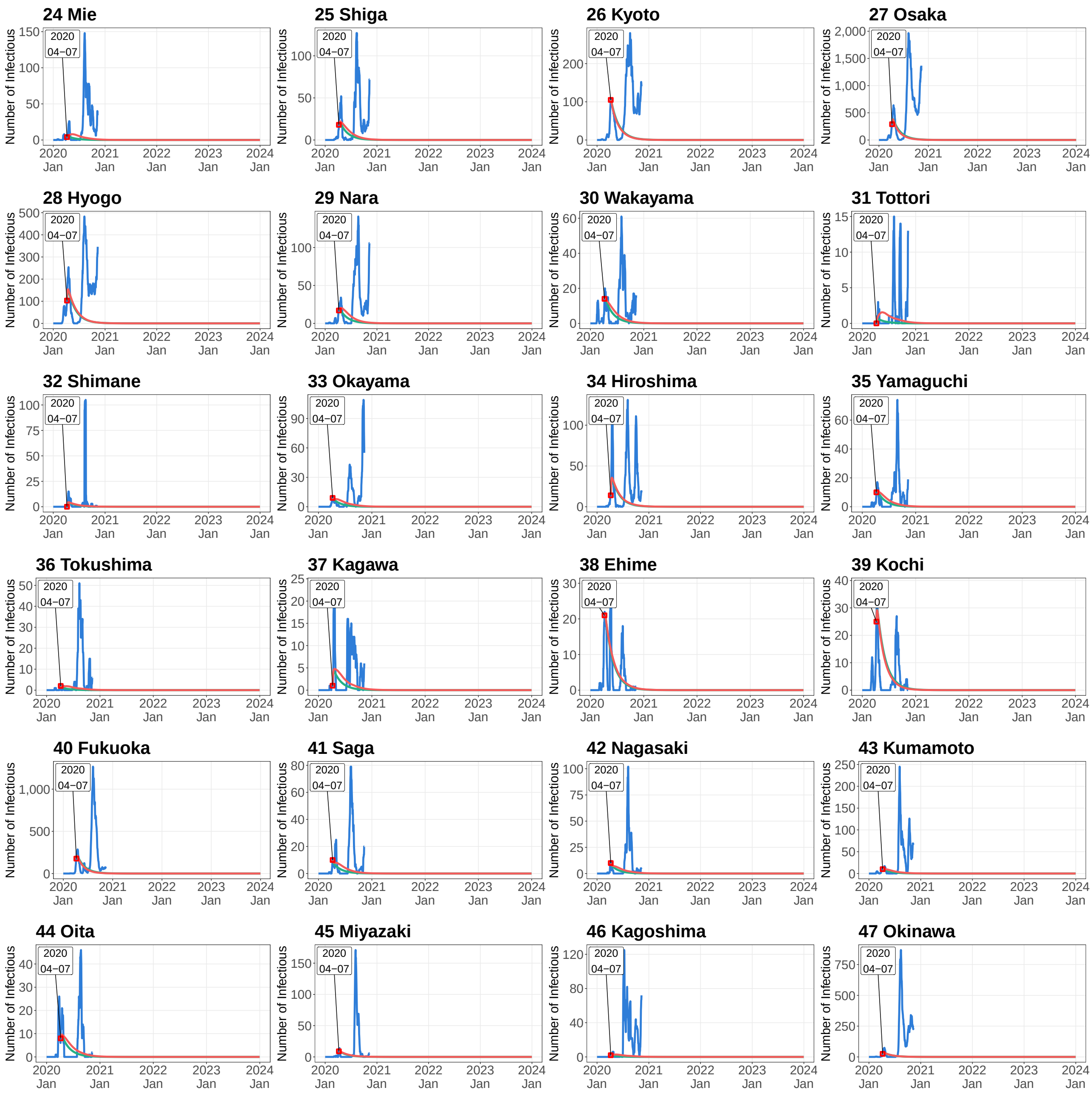


**Figure B.1.** Simulated Number of Infectious Persons by Prefecture in Case Scenario 1 (*Continued*)

Note: See caption of Figure 4 for details.

### Online Appendix C.

#### Case 2: Modest convergence case scenario with time-varying transmission rate from April 7 of 2020

Figure C.1 presents simulation results for the modest convergence case scenario with the time-varying transmission rate. The starting date of simulation is April 7 of 2020.

Table C.1 presents the specific values of intervention degree $\alpha(t)$ used in the simulation.

[Table C.1 and Figure C.1]

**Table C.1.** Parameter Setting of Intervention Degree in Case Scenario 2

| | Month | | | | | | | | | | | |
|---|---|---|---|---|---|---|---|---|---|---|---|---|
| Year | 1 | 2 | 3 | 4 | 5 | 6 | 7 | 8 | 9 | 10 | 11 | 12 |
| | Case 2 (Starting date of simulation: April 7, 2020) | | | | | | | | | | | |
| 2020 | - | - | - | 0.56 | 0.12 | 0.48 | 0.76 | 0.48 | 0.20 | 0.44 | 0.62 | 0.56 |
| 2021 | 0.40 | 0.32 | 0.20 | 0.46 | 0.32 | 0.24 | 0.48 | 0.32 | 0.24 | 0.46 | 0.54 | 0.54 |
| 2022 | 0.40 | 0.32 | 0.20 | 0.46 | 0.32 | 0.24 | 0.48 | 0.32 | 0.24 | 0.46 | 0.54 | 0.54 |
| 2023 | 0.40 | 0.32 | 0.20 | 0.46 | 0.32 | 0.24 | 0.48 | 0.32 | 0.24 | 0.46 | 0.54 | 0.54 |

Note: See caption of Table 2 for details.

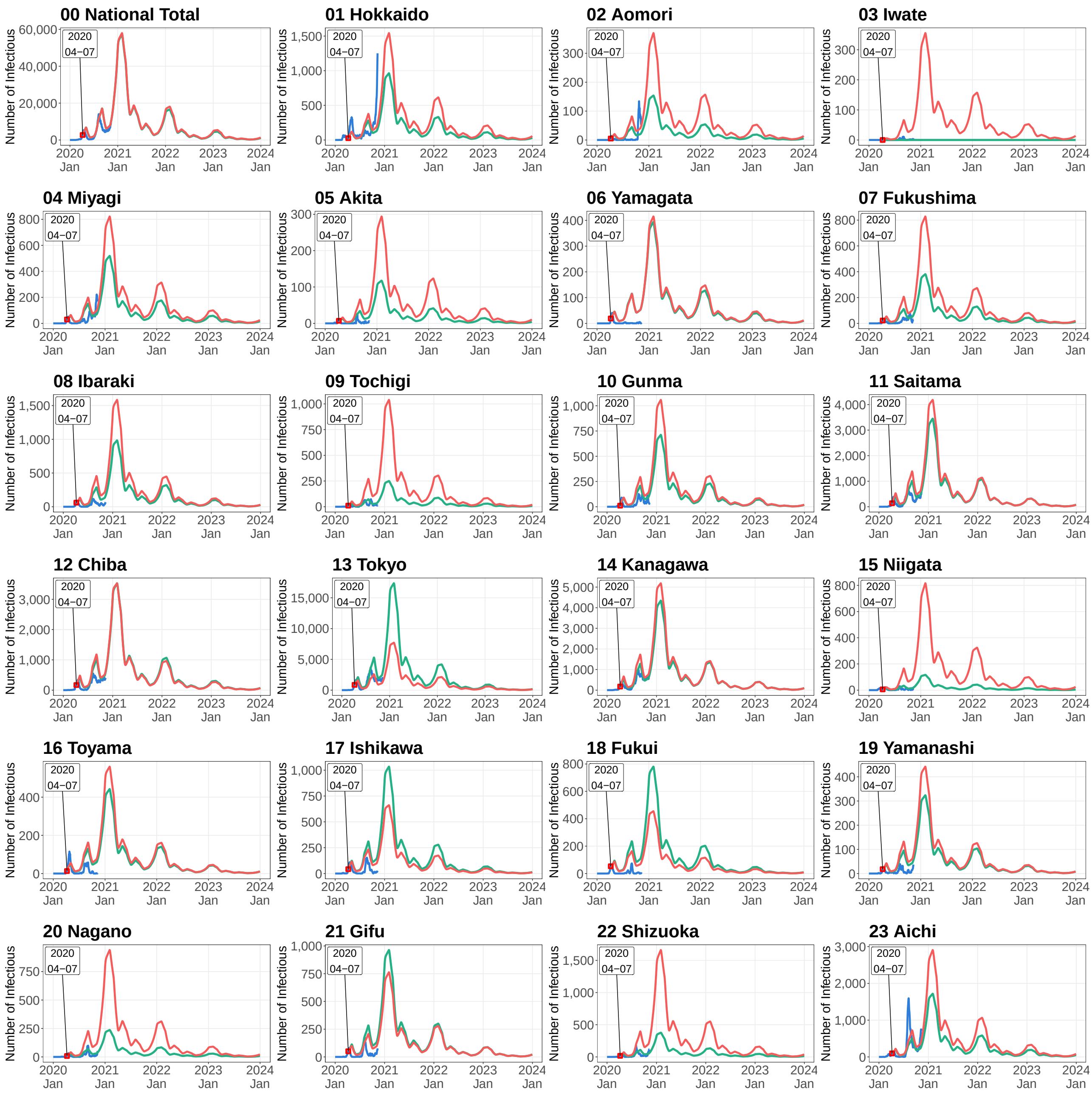


**Figure C.1.** Simulated Number of Infectious Persons by Prefecture in Case Scenario 2

Note: See caption of Figure 4 for details.

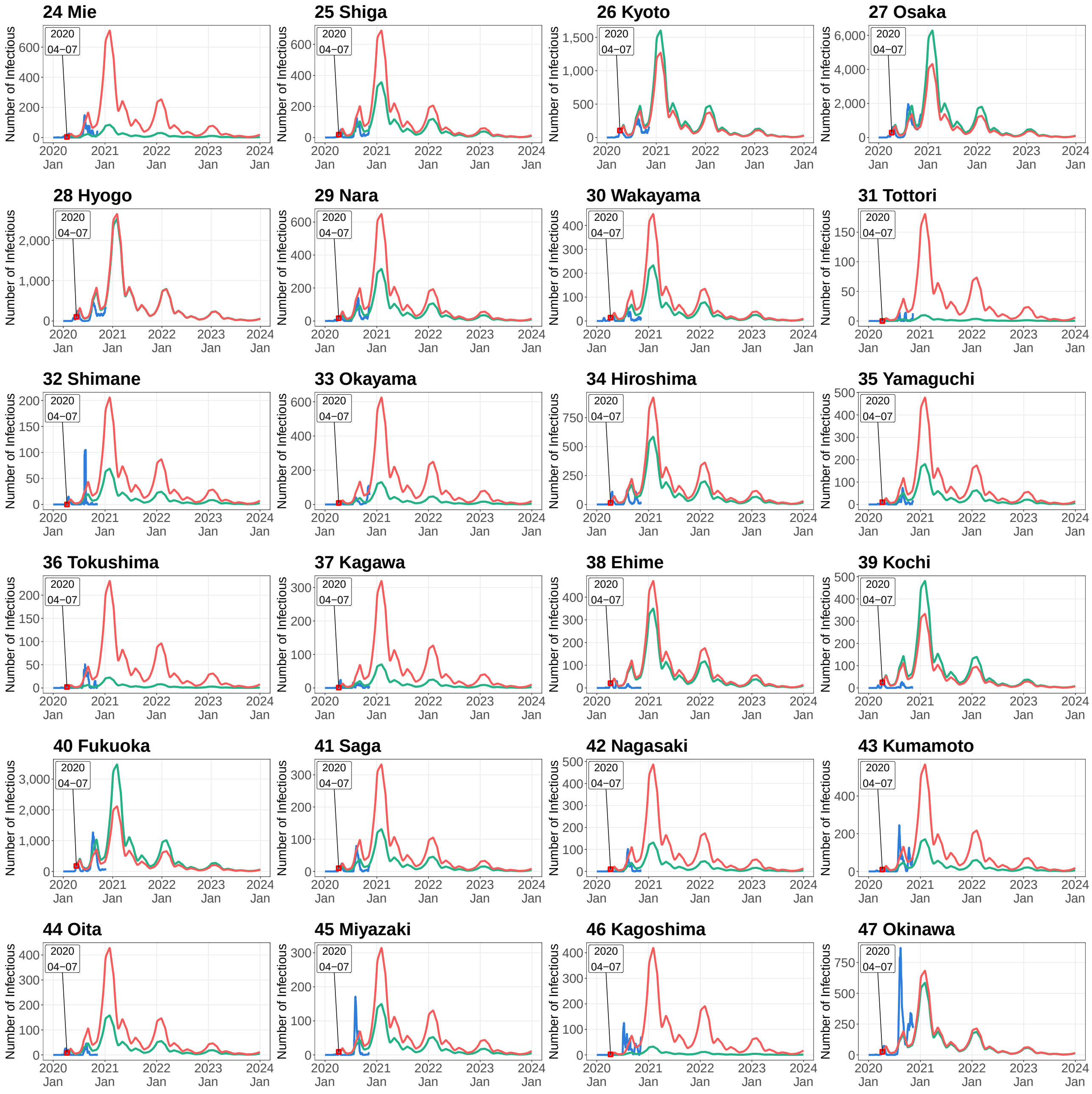


**Figure C.1.** Simulated Number of Infectious Persons by Prefecture in Case Scenario 2 (*Continued*)

Note: See caption of Figure 4 for details.

### Online Appendix D.

#### Case 3: Modest convergence case scenario with time-varying transmission rate from August 17 of 2020

Figure D.1 shows the simulation results in the modest convergence case scenario with the time-varying transmission rate. The starting date of simulation is August 17 of 2020.

Table D.1 presents the specific values of intervention degree $\alpha(t)$ used in the simulation.

[Table D.1 and D.1]

**Table D.1.** Parameter Setting of Intervention Degree in Case Scenario 3

| | Month | | | | | | | | | | | |
|---|---|---|---|---|---|---|---|---|---|---|---|---|
| Year | 1 | 2 | 3 | 4 | 5 | 6 | 7 | 8 | 9 | 10 | 11 | 12 |
| | Case 3 (Starting date of simulation: August 17 of 2020) | | | | | | | | | | | |
| 2020 | - | - | - | - | - | - | - | 0.48 | 0.20 | 0.44 | 0.62 | 0.56 |
| 2021 | 0.40 | 0.32 | 0.20 | 0.46 | 0.32 | 0.24 | 0.48 | 0.32 | 0.24 | 0.46 | 0.54 | 0.54 |
| 2022 | 0.40 | 0.32 | 0.20 | 0.46 | 0.32 | 0.24 | 0.48 | 0.32 | 0.24 | 0.46 | 0.54 | 0.54 |
| 2023 | 0.40 | 0.32 | 0.20 | 0.46 | 0.32 | 0.24 | 0.48 | 0.32 | 0.24 | 0.46 | 0.54 | 0.54 |

Note: See caption of Table 2 for details.

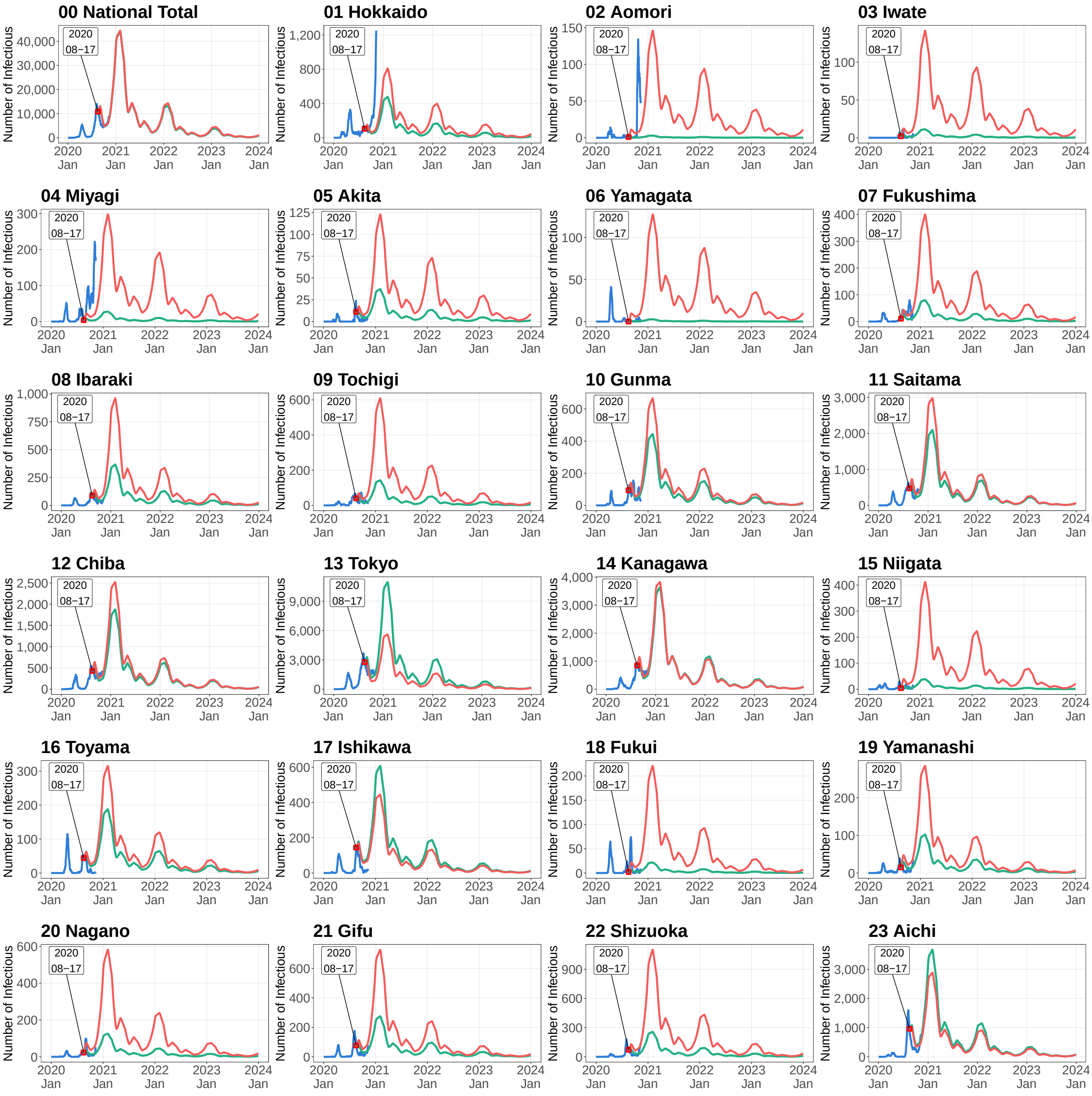


**Figure D.1.** Simulated Number of Infectious Persons by Prefecture in Case Scenario 3

Note: See caption of Figure 4 for details.

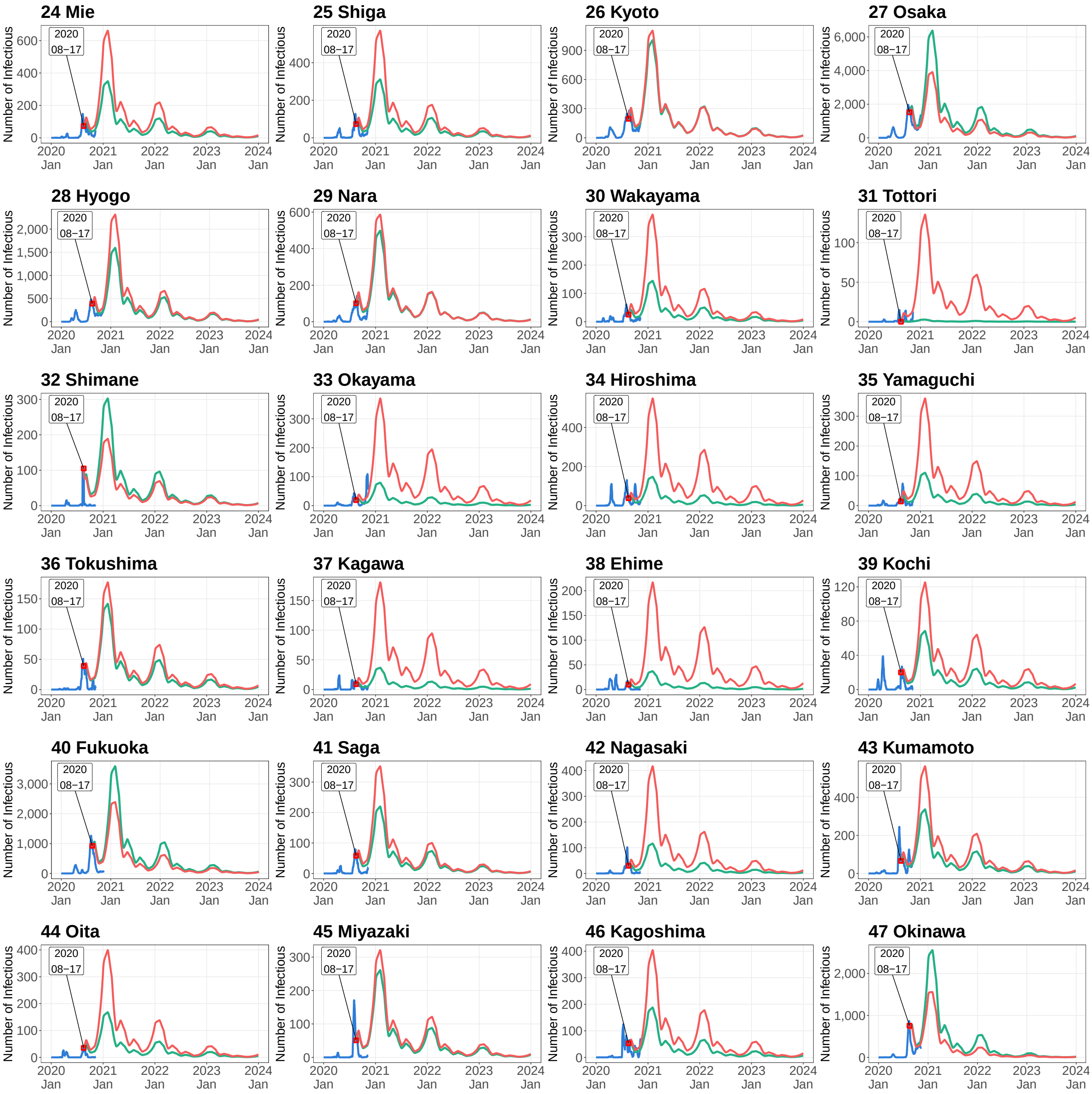


**Figure D.1.** Simulated Number of Infectious Persons by Prefecture in Case Scenario 3 (*Continued*)

Note: See caption of Figure 4 for details.

### Online Appendix E.

#### Case 4: Modest convergence case scenario with time-varying transmission rate from November 4 of 2020

Figure E.1 shows the simulation results in the modest convergence case scenario with the time-varying transmission rate. The starting date of simulation is November 4 of 2020.

Table E.1 presents the specific values of intervention degree $\alpha(t)$ used in the simulation.

[Table E.1 and E.1]

**Table E.1.** Parameter Setting of Intervention Degree in Case Scenario 4

| | Month | | | | | | | | | | | |
|---|---|---|---|---|---|---|---|---|---|---|---|---|
| Year | 1 | 2 | 3 | 4 | 5 | 6 | 7 | 8 | 9 | 10 | 11 | 12 |
| | Case 4 (Starting date of simulation: November 4 of 2020) | | | | | | | | | | | |
| 2020 | - | - | - | - | - | - | - | - | - | - | 0.62 | 0.56 |
| 2021 | 0.40 | 0.32 | 0.20 | 0.46 | 0.32 | 0.24 | 0.48 | 0.32 | 0.24 | 0.46 | 0.54 | 0.54 |
| 2022 | 0.40 | 0.32 | 0.20 | 0.46 | 0.32 | 0.24 | 0.48 | 0.32 | 0.24 | 0.46 | 0.54 | 0.54 |
| 2023 | 0.40 | 0.32 | 0.20 | 0.46 | 0.32 | 0.24 | 0.48 | 0.32 | 0.24 | 0.46 | 0.54 | 0.54 |

Note: See caption of Table 2 for details.

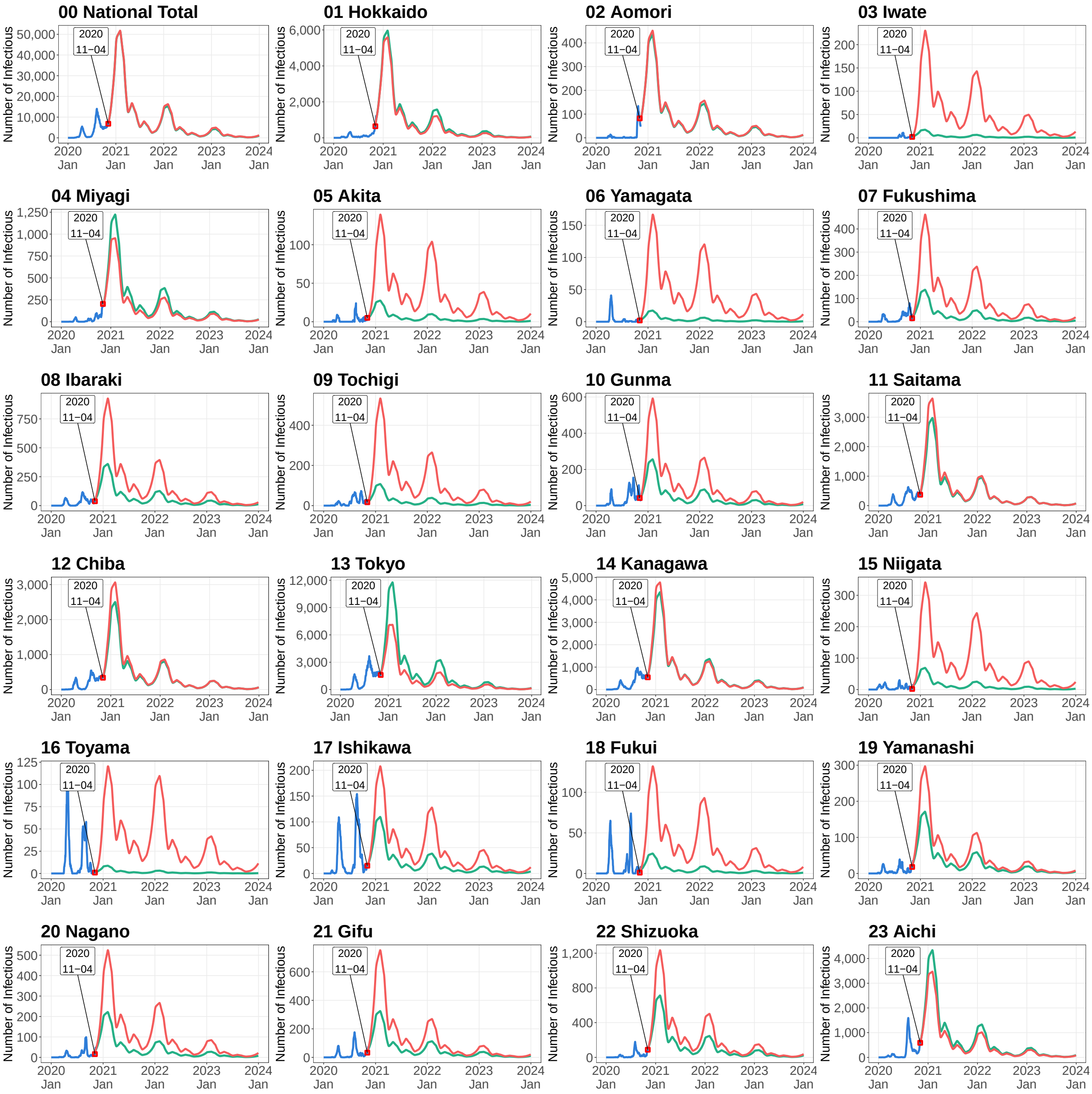


**Figure E.1.** Simulated Number of Infectious Persons by Prefecture in Case Scenario 4

Note: See caption of Figure 4 for details.

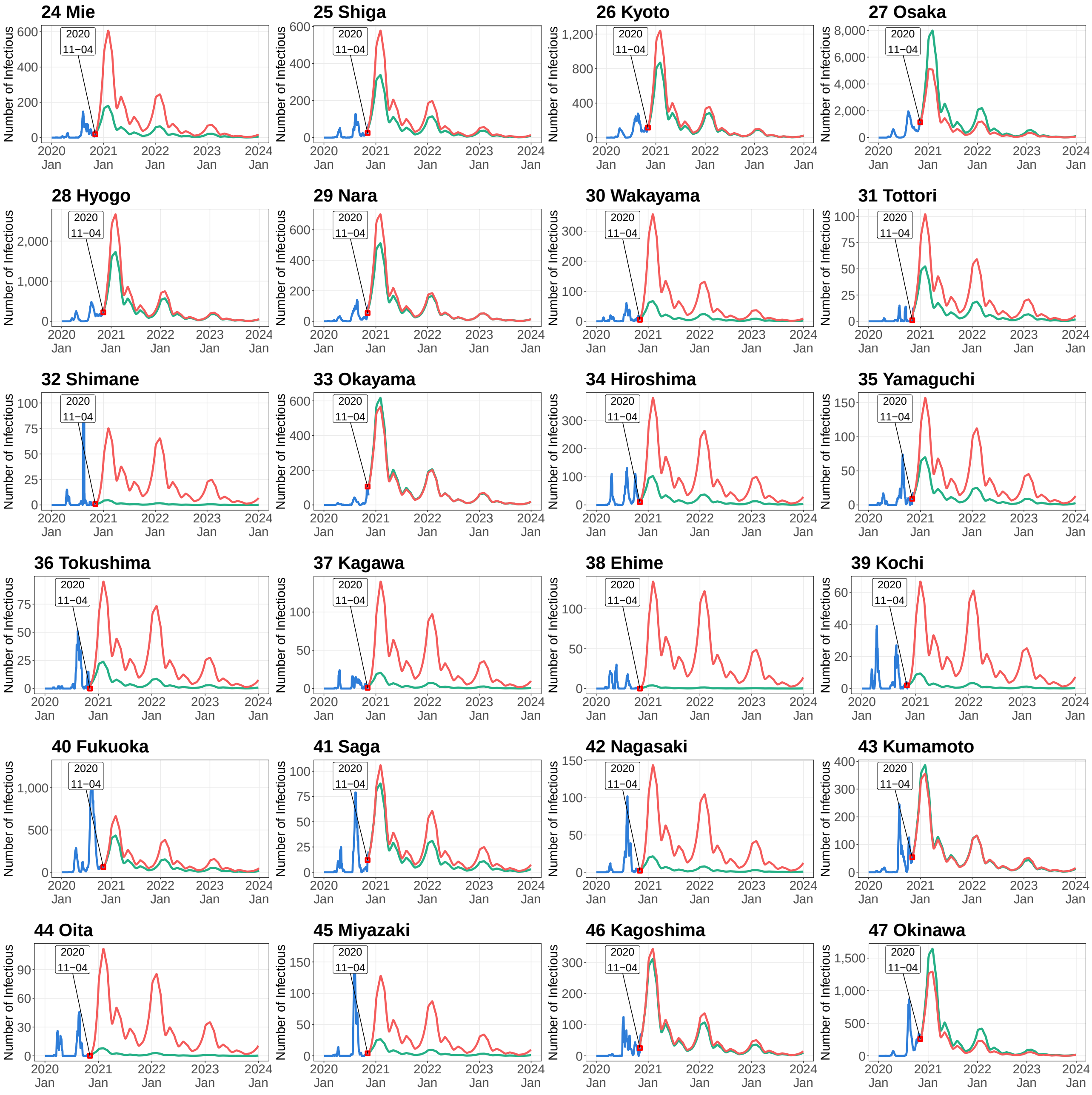


**Figure E.1.** Simulated Number of Infectious Persons by Prefecture in Case Scenario 4 (*Continued*)

Note: See caption of Figure 4 for details.

### Online Appendix F.

#### Case 5: Worsening case scenario with time-varying transmission rate from November 4 of 2020

Figure F.1 presents the simulation results in the worsening case scenario with the time-varying transmission rate. The starting date of simulation is November 4 of 2020.

Table F.1 presents the specific values of intervention degree $\alpha(t)$ used in the simulation.

[Table F.1 and Figure F.1]

**Table F.1.** Parameter Setting of Intervention Degree in Case Scenario 5

| Year | Month | | | | | | | | | | | |
|---|---|---|---|---|---|---|---|---|---|---|---|---|
| | 1 | 2 | 3 | 4 | 5 | 6 | 7 | 8 | 9 | 10 | 11 | 12 |
| | Case 5 (Starting date of simulation: November 4 of 2020) | | | | | | | | | | | |
| 2020 | - | - | - | - | - | - | - | - | - | - | 0.62 | 0.56 |
| 2021 | 0.50 | 0.46 | 0.40 | 0.34 | 0.34 | 0.32 | 0.40 | 0.36 | 0.40 | 0.44 | 0.54 | 0.54 |
| 2022 | 0.50 | 0.46 | 0.40 | 0.34 | 0.34 | 0.32 | 0.40 | 0.36 | 0.40 | 0.44 | 0.54 | 0.54 |
| 2023 | 0.50 | 0.46 | 0.40 | 0.34 | 0.34 | 0.32 | 0.40 | 0.36 | 0.40 | 0.44 | 0.54 | 0.54 |

Note: See caption of Table 2 for details.

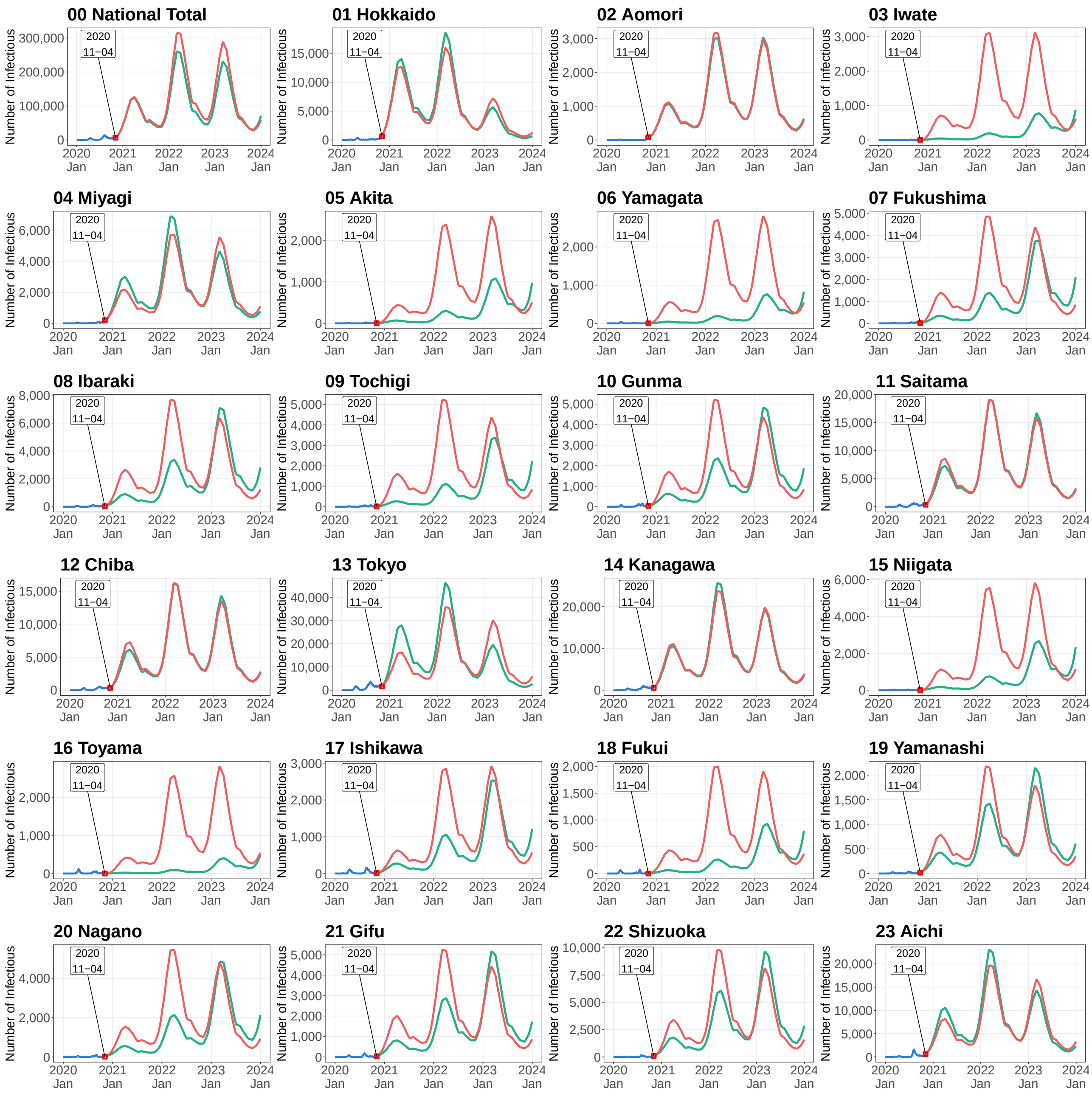


**Figure F.1.** Simulated Number of Infectious Persons by Prefecture in Case Scenario 5

Note: See caption of Figure 4 for details.

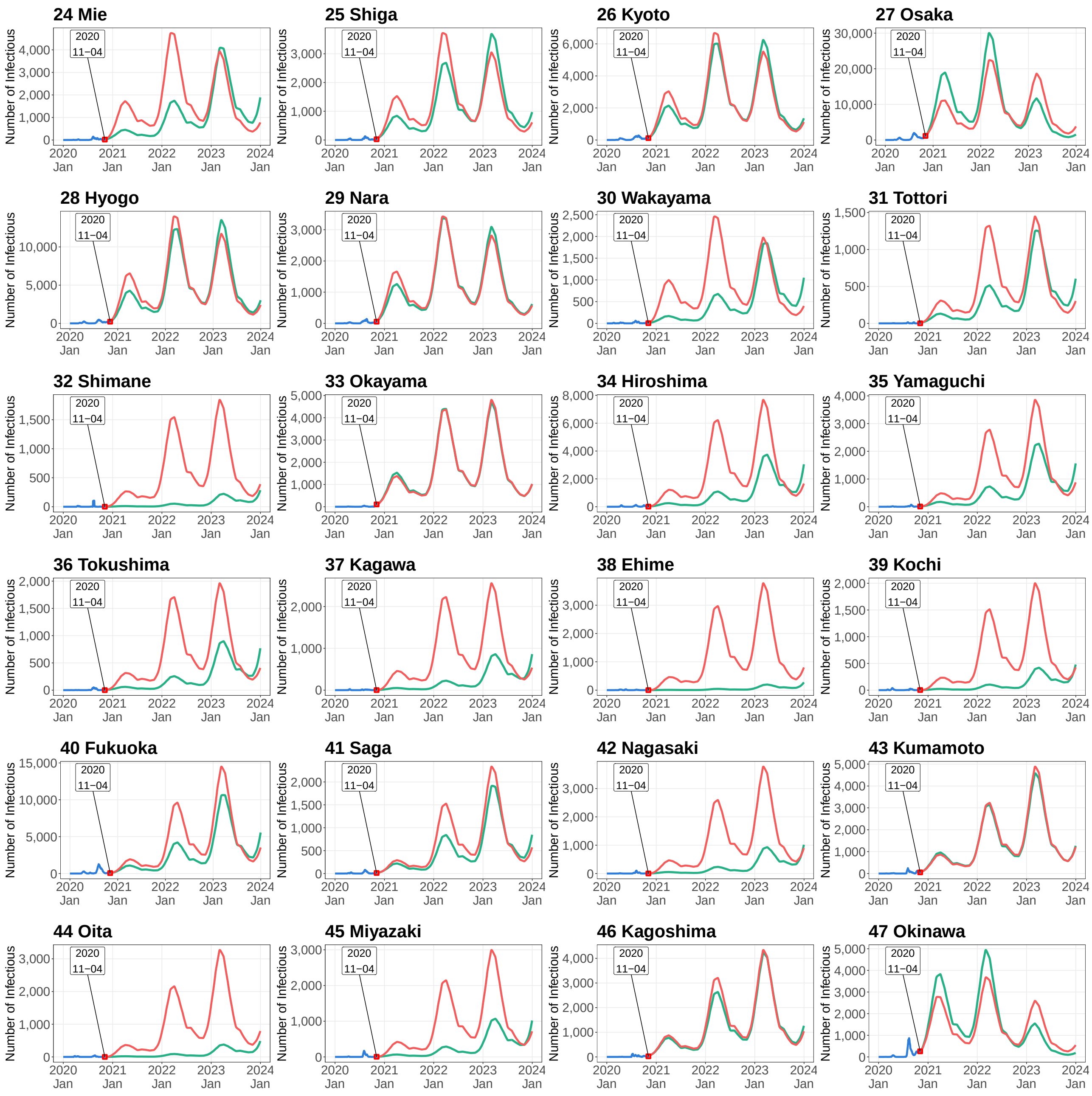


**Figure F.1.** Simulated Number of Infectious Persons by Prefecture in Case Scenario 5 (*Continued*)

Note: See caption of Figure 4 for details.

### Online Appendix G.

#### Case 6: Mobility restriction only for Tokyo and modest convergence case scenario with time-varying transmission rate from November 4 of 2020

Figure G.1 shows the simulation results under the mobility restriction only for Tokyo in the modest convergence case scenario with the time-varying transmission rate. The starting date of simulation is November 4 of 2020.

Table G.1 presents the specific values of intervention degree $\alpha(t)$ used in the simulation.

[Table G.1 and Figure G.1]

**Table G.1.** Parameter Setting of Intervention Degree in Case Scenario 6

| | Month | | | | | | | | | | | |
|---|---|---|---|---|---|---|---|---|---|---|---|---|
| Year | 1 | 2 | 3 | 4 | 5 | 6 | 7 | 8 | 9 | 10 | 11 | 12 |
| | Case 6 (Starting date of simulation: November 4 of 2020) | | | | | | | | | | | |
| 2020 | - | - | - | - | - | - | - | - | - | - | 0.62 | 0.56 |
| 2021 | 0.50 | 0.46 | 0.40 | 0.34 | 0.34 | 0.32 | 0.40 | 0.36 | 0.40 | 0.44 | 0.54 | 0.54 |
| 2022 | 0.50 | 0.46 | 0.40 | 0.34 | 0.34 | 0.32 | 0.40 | 0.36 | 0.40 | 0.44 | 0.54 | 0.54 |
| 2023 | 0.50 | 0.46 | 0.40 | 0.34 | 0.34 | 0.32 | 0.40 | 0.36 | 0.40 | 0.44 | 0.54 | 0.54 |

Note: See caption of Figure 4 for details.

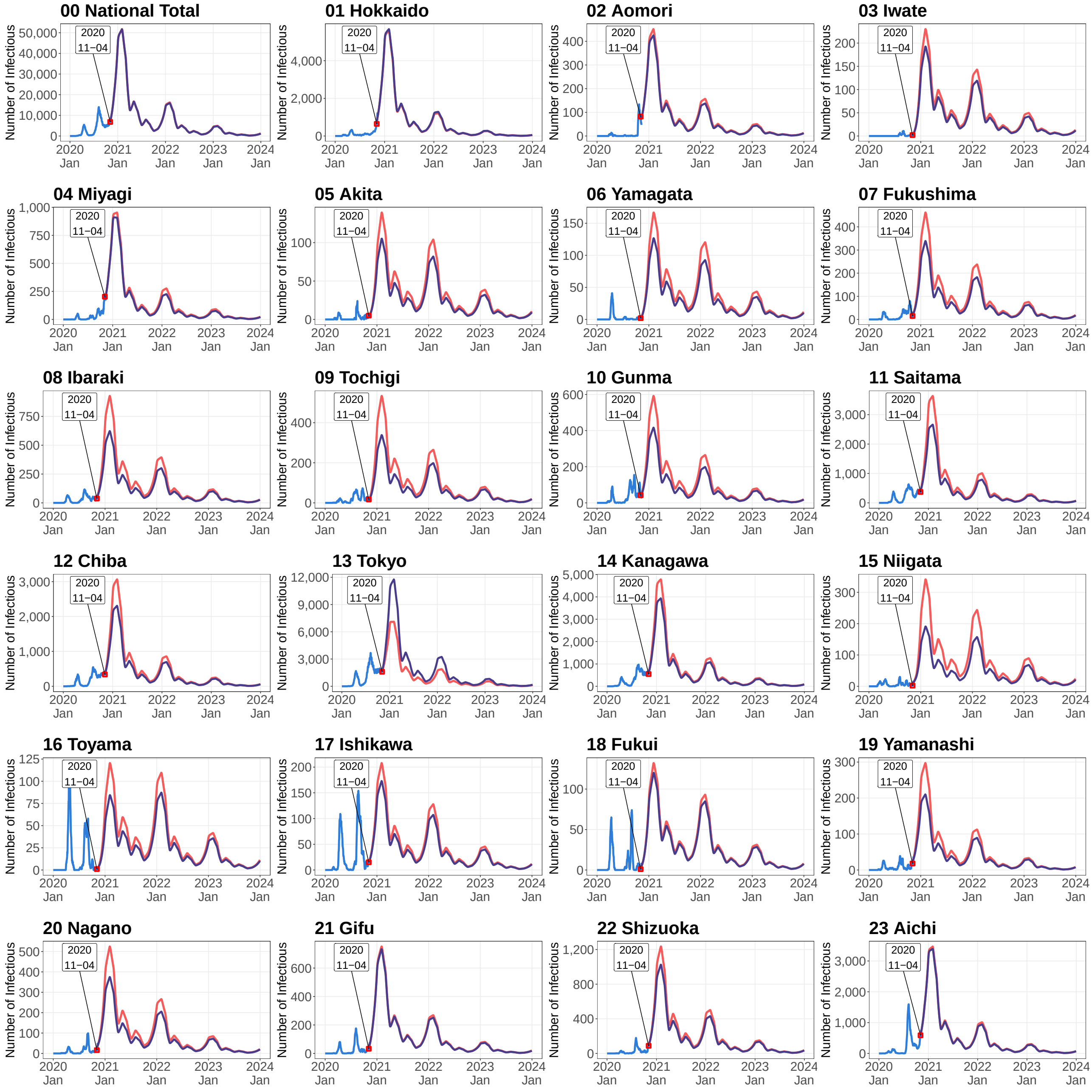


**Figure G.1.** Simulated Number of Infectious Persons by Prefecture in Case Scenario 6

Note: Shown are the observed numbers of infectious individuals (blue lines), and the numbers of infectious individuals simulated by the spatial SEIR model with interregional mobility and with interregional mobility except Tokyo (red and purple lines, respectively). Simulations were started on November 4 of 2020.

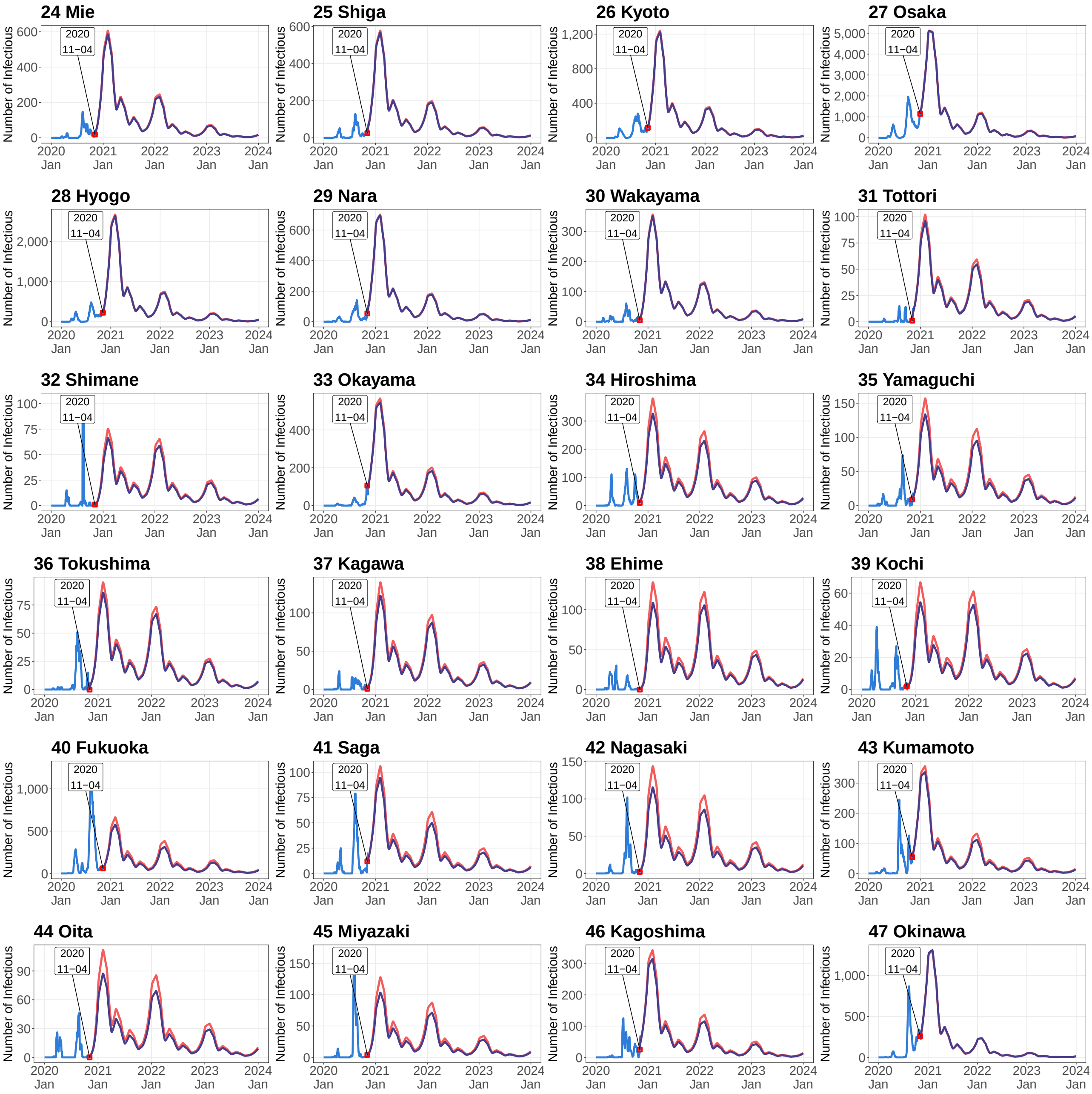


**Figure G.1.** Simulated Number of Infectious Persons by Prefecture in Case Scenario 6 (*Continued*)

Note: See caption of Figure G.1 for details.
